# A unified deep learning framework for cross-platform harmonization of multi-tracer PET quantification

**DOI:** 10.1101/2025.10.20.25338339

**Authors:** Jing Wang, Aocheng Zhong, Qian Xu, Haolin Huang, Yuhua Zhu, Jiayin Lu, Min Wang, Jiehui Jiang, Chengyang Li, Ming Ni, Kaicong Sun, Yihui Guan, Jie Lu, Mei Tian, Dinggang Shen, Huiwei Zhang, Qian Wang, Chuantao Zuo

## Abstract

Quantitative PET underpins diagnosis and treatment monitoring in neurodegenerative disease, yet systematic biases between PET-MRI and PET-CT preclude threshold transfer and cross-site comparability. We present a unified, anatomically guided deep-learning framework that harmonizes multi-tracer PET-MRI to PET-CT. The model learns CT-anchored attenuation representations with a Vision Transformer Autoencoder, aligns MRI features to CT space via contrastive objectives, and performs attention-guided residual correction. In paired same-day scans (*N* = 70; amyloid, tau, FDG), cross-platform bias fell by >80% while preserving inter-regional biological topology. The framework generalized zero-shot to held-out tracers (^18^F-florbetapir; ^18^F-FP-CIT) without retraining. Multicentre validation (*N* = 420; three sites, four vendors) reduced amyloid Centiloid discrepancies from 23.6 to 4.1 (within PET-CT test-retest precision) and aligned tau SUVR thresholds. These results enable platform-agnostic diagnostic cutoffs and reliable longitudinal monitoring when patients transition between modalities, establishing a practical route to scalable, radiation-sparing quantitative PET in therapeutic workflows.

## Introduction

Hybrid PET-MRI enables simultaneous molecular and anatomical imaging with substantially reduced radiation exposure compared with PET-CT^1–3^. With disease-modifying therapies entering clinical practice^4,5^, quantitative PET now plays critical theranostic roles in patient selection and treatment monitoring for AD and other neurodegenerative disorders^6,7^. Current clinical criteria explicitly require high quantitative fidelity for amyloid and tau PET^6,8^. However, platform-dependent measurement variability between PET-MRI and PET-CT prevents direct threshold transfer and cross-site comparability. Although standardization frameworks such as the Centiloid scale (CL)^9–12^ for amyloid and CenTauR scale^13^ for tau established on PET-CT have improved interpretability across tracers and pipelines, they do not eliminate PET-MRI-specific biases^9,14^. These platform effects can confound diagnostic decisions near clinical cutoffs and reduce sensitivity to longitudinal change, both of which are critical for monitoring treatment response^15–17^.

Quantification inconsistencies in PET-MRI arise from intrinsic physical differences and platform-dependent variability. A key source is attenuation correction (AC): CT directly measures tissue attenuation, whereas MRI must estimate it indirectly from proton density and relaxation signals^18,19^. This makes MRI-based AC inherently more challenging and sensitive to protocol choices, introducing >10% bias for amyloid and tau tracers^20–23^. Even with state-of-the-art AC methods, multi-centre studies report 10–25% quantitative discrepancies across platforms^24,25^, which are significant for disease staging and treatment monitoring^15–17^. Existing harmonization strategies, such as EARL and vendor-specific calibrations^25,26^, improve cross-site consistency but remain limited by tracer-specific tuning, vendor dependencies, and susceptibility to regional artifacts^20,21,27–29^. As new radiotracers emerge for synaptic density, neuroinflammation, and other targets, scalable and tracer-agnostic harmonization becomes essential for multi-tracer imaging studies.

To address the quantification inconsistencies in PET-MR, recent advances in deep learning offer potential solutions. Sophisticated network architectures such Vision Transformer (ViT) have demonstrated strong capabilities in capturing long-range anatomical dependencies critical for accurate attenuation modeling^30–32^ and multi-modal learning^33,34^. Generative Adversarial Network (GANs) enable cross-modality translation through its strong capability in learning and mapping data distributions^35^, while autoencoders have emerged as powerful self-supervised learning tool for robust image representations^36^. However, despite these technological advances^37,38^, no unified framework exists for multi-tracer PET harmonization that generalizes across radiotracers and scanner platforms while preserving biologically meaningful signal patterns.

Here we present a unified deep learning framework that harmonizes PET-MRI quantification to PET-CT across tracers and vendors. The framework integrates three components: (i) CT-anchored anatomical representation learning, (ii) MRI-to-CT feature alignment via contrastive learning, and (iii) attention-guided residual PET correction. Multi-modal alignment of MRI and CT representations enhance cross-domain alignment of PET-MRI and PET-CT. Trained on same-day paired acquisitions (*N* = 70; amyloid, tau, FDG), reduced cross-platform bias by >80% while preserving inter-regional correlation patterns and rank ordering of regional uptake values. Without retraining, the framework generalized to unseen tracers (^18^F-florbetapir, ^18^F-FP-CIT) with comparable harmonization performance, demonstrating tracer-agnostic capability. Multi-centre validation (*N* = 420; three sites, four vendors) showed that harmonized PET-MRI amyloid CL values aligned with PET-CT within test-retest variability, reducing discrepancy from 23.6 to 4.1 CL units. These results enable platform-agnostic diagnostic thresholds and seamless longitudinal monitoring when patients transition between modalities, establishing a practical foundation for radiation-sparing quantitative PET in clinical workflows^4–8,10–14^.

## Results

### Benchmarking harmonized whole-image quality

We evaluated harmonization performance using same-day paired PET-CT and PET-MRI acquisitions from 70 participants across three tracers: ^18^F-florbetaben (amyloid, *N* = 20), ^18^F-florzolotau (tau, *N* = 30), and ^18^F-FDG (glucose metabolism, *N* = 20) (Fig.1, Extended Data Table 1). The inter-scan interval (5–7 min) minimized tracer redistribution, ensuring PET-CT could serve as ground truth for quantitative benchmarking.

**Fig. 1.**
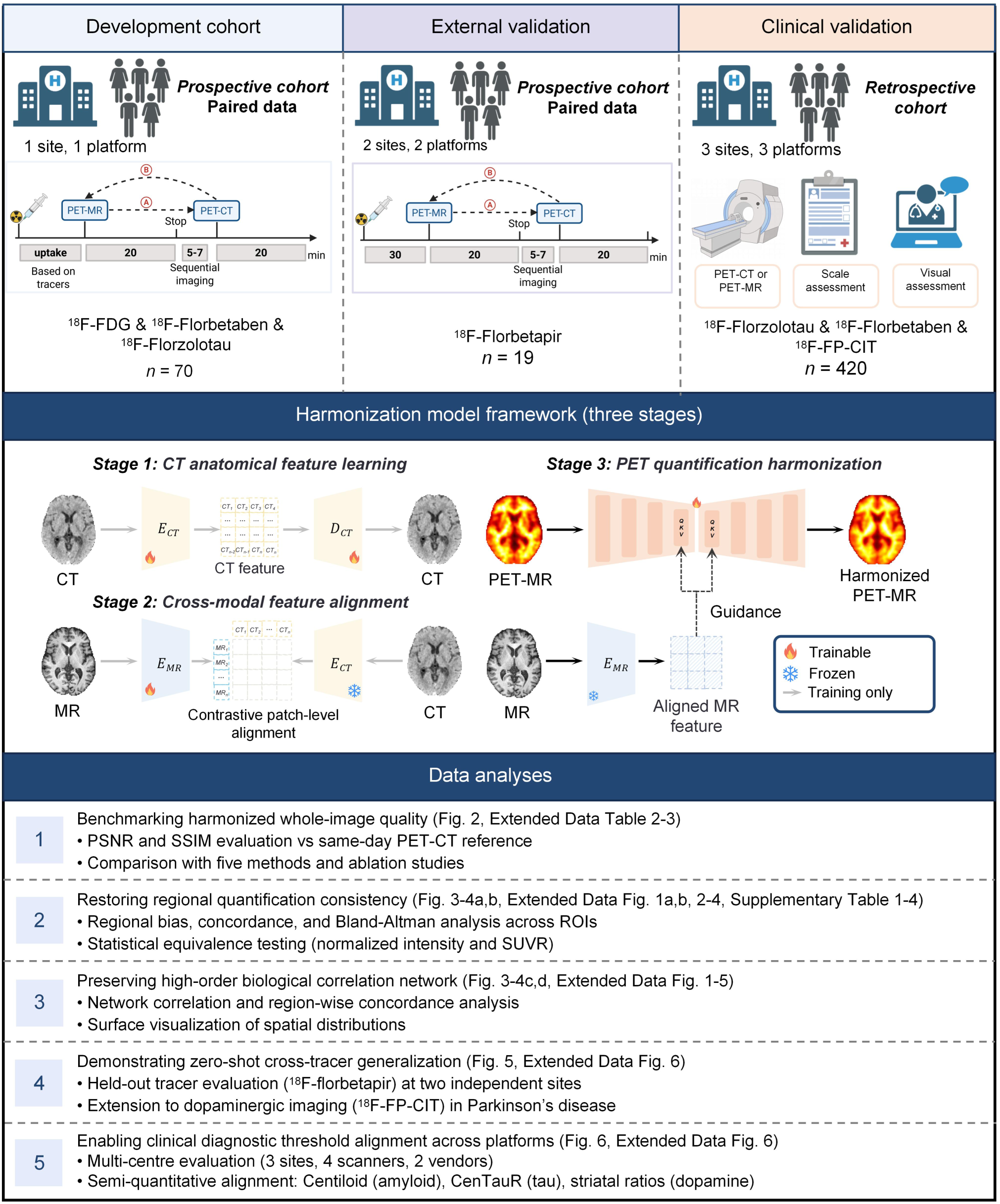
Study design and framework overview. **Upper**: Three-phase study. *Development* (*N* = 70): prospective same–day paired PET–CT and PET–MRI across ^18^F–FDG (*N* = 20), ^18^F–florbetaben (*N* = 20) and ^18^F–florzolotau (*N* = 30); inter–scan interval 5–7 min to minimise redistribution. *External validation* (*N* = 19): cross–vendor evaluation on ^18^F–florbetapir (held–out tracer) at two independent centres. *Clinical validation* (*N* = 420): multicentre retrospective evaluation spanning three sites and four vendor configurations. **Middle**: Three–stage framework. *Stage 1*, CT–based anatomical feature extraction (Vision Transformer Autoencoder). *Stage 2*, cross–modal MRI→CT alignment via contrastive learning. *Stage 3*, anatomically guided residual PET correction using aligned MR features. Trainable modules (fire icon); frozen components (snowflake). **Lower**: Evaluation schema: (1) image–quality benchmarking (PSNR, SSIM), (2) regional quantification with paired comparisons, regression and Bland-Altman agreement, (3) cross–tracer generalization (held–out ^18^F–florbetapir; extension to ^18^F–FP–CIT), and (4) clinical validation (amyloid CL cut–offs; tau CenTauR meta–ROI SUVR).

Our framework outperformed five established methods across all tracers (unharmonized PET-MRI, SwinIR^39^, Restormer^40^, DRMC^41^, AIMIR^42^) (Fig.2, Extended Data Table 2). For ^18^F-florbetaben, peak signal-to-noise ratio (PSNR) improved from 31.41 dB (unharmonized PET-MRI) to 34.80 ± 2.13 dB and structural similarity index (SSIM) from 0.97 to 0.98 ± 0.005. Similar improvements were observed for ^18^F-florzolotau (PSNR: 34.54 to 36.77 dB; SSIM: 0.97 to 0.98) and ^18^F-FDG (PSNR: 36.18 to 37.25 dB; SSIM: 0.98 to 0.99), indicating effective artifact suppression while preserving anatomical detail. Ablation studies confirmed that all architectural components (structural guidance, MR-to-CT alignment) contributed meaningfully to this performance (Extended Data Table 3). These metrics validate the framework’s ability to deliver harmonized images suitable for PET-CT-grade quantification, establishing a foundation for quantitative biomarker extraction.

**Fig. 2.**
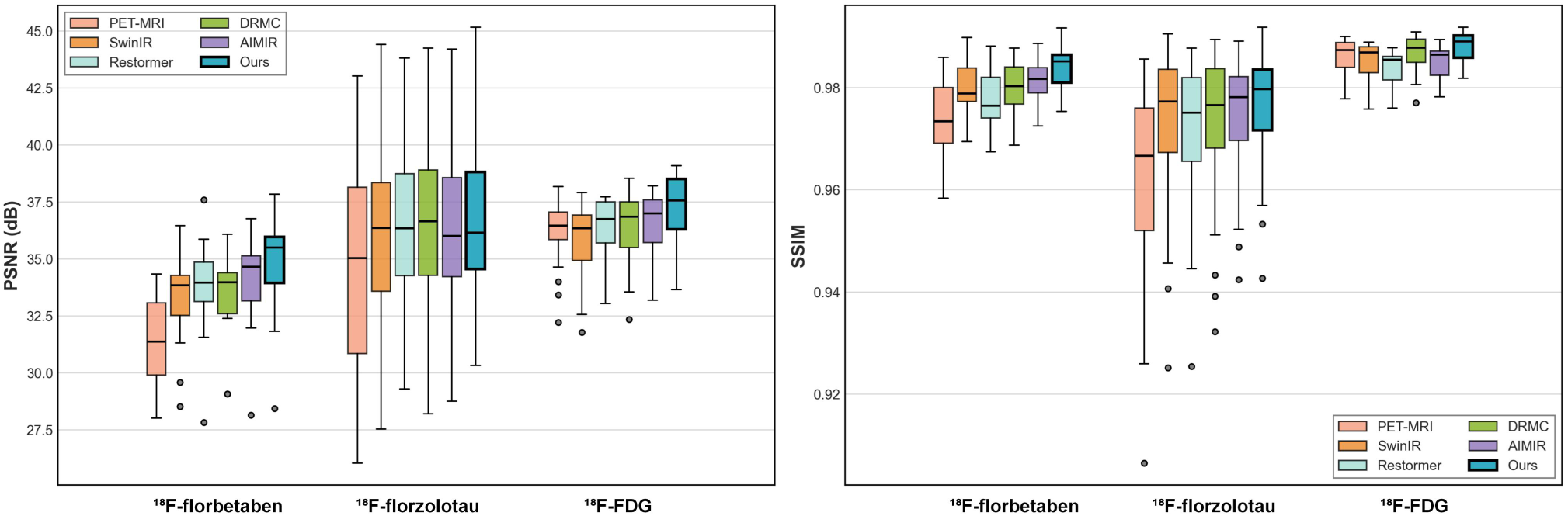
Quantitative benchmarking of harmonization performance. Peak signal-to-noise ratio (PSNR) and structural similarity index (SSIM) for three tracers harmonized by six methods. Box plots: center line, median; box, IQR; whiskers, 1.5× IQR; points, individual participants. Our framework achieved (mean ± s.d.): ^18^F-florbetaben, PSNR 34.80 ± 2.13 dB, SSIM 0.98 ± 0.005; ^18^F-florzolotau, PSNR 36.77 ± 3.67 dB, SSIM 0.98 ± 0.011; ^18^F-FDG, PSNR 37.25 ± 1.58 dB, SSIM 0.99 ± 0.003. Data from five-fold cross-validation (*N* = 70 total: ^18^F-florbetaben *N* = 20, ^18^F-florzolotau *N* = 30, ^18^F-FDG *N* = 20). Methods compared: unharmonized PET-MRI, SwinIR^39^, Restormer^40^, DRMC^41^, AIMIR^42^, and our framework. See Extended Data Table 2 for detailed metrics and statistical comparisons.

### Restoring regional quantification consistency

We quantified regional platform bias using percent bias, cross-platform concordance, and Bland-Altman analysis across three tracers (Fig.3-4a,b, Extended Data Fig.1a,b, 2-4, Supplementary Table 5). Harmonization reduced regional biases to <1% for amyloid and <5% for tau, achieving statistical equivalence with PET-CT (all *P* > 0.05, Bonferroni-corrected). Cross-platform concordance remained high (*r* > 0.85) across all tracers and regions, demonstrating preservation of inter-subject variability essential for diagnostic classification.

**Fig. 3.**
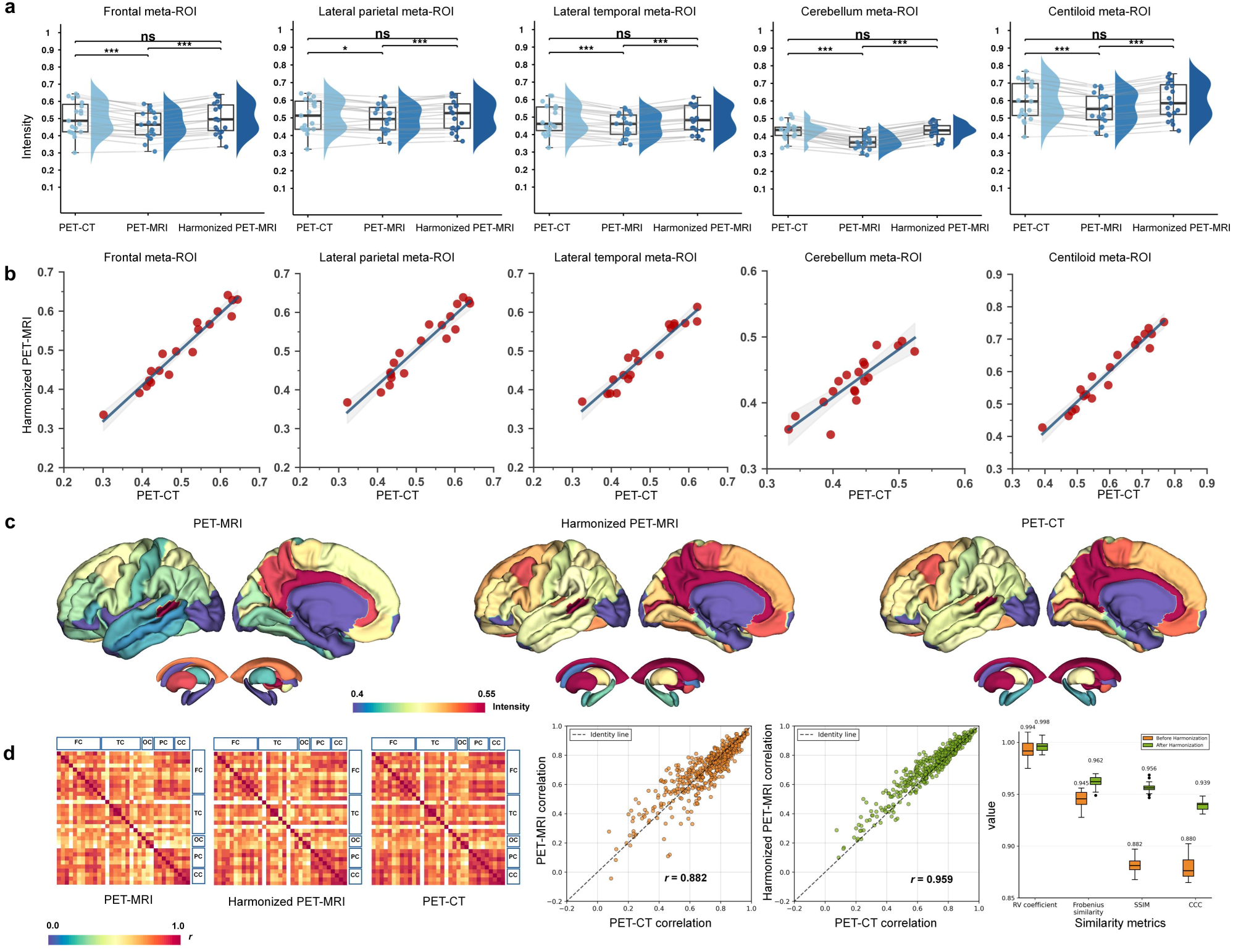
Cross-platform harmonization of ^18^F-florbetaben quantification. **a**, Regional normalized intensity distributions comparing PET-CT (reference), unharmonized PET-MRI, and harmonized PET-MRI across five brain regions. Violin plots: median (center line) and quartiles. Statistical comparisons: Wilcoxon signed-rank test with Bonferroni correction. \*\*\**P* < 0.001; \*\**P* < 0.01; \**P* < 0.05; *ns* not significant (*P* > 0.05). Harmonization achieved statistical equivalence with PET-CT across all regions. **b**, Cross-platform concordance for five regions. Each panel: scatter plot (harmonized PET-MRI versus PET-CT), regression line, Pearson correlation coefficient (*r*), coefficient of determination (*R*^2^), and *P* value. Near-unity slopes (0.73–0.93) and high correlations (*r* > 0.85, all *P* < 0.001) demonstrate preserved inter-subject variability. See Extended Data Table 5 for detailed regression parameters. **c**, Cortical surface projections showing preserved amyloid distribution patterns. Color scale: normalized intensity (0.4–0.55). **d**, Inter-regional correlation matrices (33 regions). Left: correlation matrices for unharmonized and harmonized PET-MRI versus PET-CT. Middle: matrix-level correlation improvement (*r* = 0.88 to 0.96, *P* < 0.001). Right: network similarity metrics. Region-pair analysis (Extended Data Fig. 5a) shows 97% of regions with improved concordance.

For ^18^F-florbetaben, unharmonized PET-MRI exhibited regional biases ranging from -16.18% (cerebellum) to -3.13% (lateral parietal cortex) relative to PET-CT (all *P* < 0.05, Bonferroni-corrected; Fig.3a-b). Harmonization reduced all regional biases to <1% (range: 0.10–0.70%) and decreased Centiloid meta-ROI bias from -7.06% to 0.39% (94% reduction), achieving statistical equivalence with PET-CT (all *P* > 0.05). Cross-platform concordance was excellent (*r* = 0.97, slope = 0.93, *P* < 0.001; Fig. 3b), with Bland–Altman mean bias improving from −8.3% to 0.5% and limits of agreement (LoA) narrowing from [-22.7%, 6.2%] to [-9.5%, 10.4%] (Extended Data Fig.3a). These results demonstrate correction of systematic bias while preserving inter-subject variability essential for amyloid classification.

For ^18^F-florzolotau, baseline platform bias was larger, ranging from -21.61 ± 10.03% (cerebellar gray matter) to -15.01% (lateral temporal cortex) (Fig.4a). Harmonization achieved substantial reductions: cerebellar gray matter bias decreased by 79% (to -4.57 ± 13.30%), frontal meta-ROI by 72% (to -3.85%), and CenTauR composite by 59% (to -4.71%) (all *P* < 0.001). Cross-platform concordance for the CenTauR meta-ROI was excellent (*r* = 0.96, slope = 0.96, *P* < 0.001; Fig.4b), preserving spatial patterns critical for Braak staging. Bland–Altman analysis showed pooled mean bias reduced from -14.0% to -4.7%, with LoA narrowing from [-35.3%, 7.2%] to [-25.8%, 16.4%] (Extended Data Fig.3b). These improvements demonstrate effective platform harmonization despite challenging baseline conditions.

For ^18^F-FDG, baseline platform differences were minimal (bias range: -3.47% to +2.73%), likely reflecting mature vendor calibration protocols (Extended Data Fig.1a). Harmonization further refined quantification (biases: -0.72% to 0.92%) while maintaining strong cross-platform concordance (*r* = 0.85–0.92, *P* < 0.001; Extended Data Fig.1b). Bland-Altman analysis showed pooled mean bias reduced from -1.2% to -0.1% (92% reduction), with LoA narrowing from [-12.3%, 10.0%] to [-8.4%, 8.1%] (Extended Data Fig.3c), demonstrating enhanced precision for both systematic and random error components.

Parallel analyses using standardized uptake value ratios (SUVR) confirmed consistent bias reduction across all tracers (Extended Data Fig.2,4, Supplementary Tables 2-4), demonstrating that harmonization effects were independent of the quantification metric. Residual cross-platform disagreement fell within clinically accepted bounds, enabling direct transfer of PET-CT-derived diagnostic thresholds to PET-MRI.

### Preserving high-order biological correlation network

Beyond global and regional bias correction, we assessed whether harmonization preserved disease-relevant inter-regional uptake patterns (Fig.3d and 4d; Extended Data Fig.5). For ^18^F-florbetaben, correlation between the inter-regional networks of harmonized PET-MRI and PET-CT improved from *r* = 0.88 to *r* = 0.96 (*P* < 0.001), with convergent improvements across multiple network metrics. Among 33 brain regions, 97% showed enhanced concordance, with the largest gains in cortical regions critical for amyloid staging (Extended Data Fig.5a). Similar improvements were observed for ^18^F-florzolotau (network correlation: 0.98 to 0.99, 91% regions improved) and ^18^F-FDG (0.96 to 0.97, 79% regions improved), with notable gains in regions following known disease pathways (Extended Data Fig.5b-c). Cortical surface projections confirmed preservation of tracer-specific spatial distributions (Fig.3c and 4c; Extended Data Fig.1c).

These results demonstrate that harmonization corrects systematic bias while maintaining the biological signal topology required for pathological staging and network-level disease characterization.

### Demonstrating zero-shot cross-tracer generalization

To evaluate generalization beyond training data, we tested the framework on ^18^F-florbetapir, a tracer unseen during training, using two independent centers and platforms (Siemens and United Imaging) (Fig.5). Unharmonized PET-MRI exhibited significant platform bias across all composite regions (all *P* < 0.05, Wilcoxon signed-rank test). Without retraining, zero-shot harmonization achieved statistical equivalence with PET-CT (all *P* > 0.05) while preserving inter-subject variability (correlation with PET-CT: *r* > 0.77, all *P* < 0.001).

Cortical surface projections demonstrated that harmonized PET-MRI faithfully reproduced characteristic amyloid distribution patterns observed in PET-CT at both sites (Fig.5a-b), indicating physics-driven correction rather than tracer-specific fitting. Consistent performance across an unseen tracer and multiple vendors (*N* = 19 total: Site 1 *N* = 7, Site 2 *N* = 12) demonstrates that the framework captures generalizable platform differences, supporting deployment without site- or tracer-specific recalibration.

### Enabling clinical diagnostic threshold alignment across platforms and tracers

A critical clinical barrier to PET-MRI adoption is the inability to directly apply PET-CT-derived diagnostic thresholds. We validated threshold transferability in 420 participants across three sites with four scanner configurations spanning platforms from two vendors (Fig.6, Extended Data Fig. 6). For ^18^F-florbetaben, multi-centre validation (three sites, four scanners) demonstrated successful threshold alignment. PET-CT yielded an optimal CL threshold of 22.6 (consistent with established ranges of 25–35)^6,10,43^, whereas unharmonized PET-MRI required 46.2, representing a 104% discrepancy. Harmonization reduced the threshold to 26.7 (83% bias reduction), achieving agreement within clinically acceptable limits. Harmonized values showed excellent correlation with PET-CT (*r* = 0.99, slope = 0.92, RMSE = 4.95 CL; Fig.6a-b). Critically, harmonization reduced cross-platform discrepancies from 23.6 to 4.1 CL, which approaches the estimated test–retest variability of PET-CT (∼3 CL)^9^ and represents <10% of treatment effects observed with lecanemab (−59 units over 18 months)^4^. This precision enables reliable longitudinal monitoring when patients transition between platforms during therapy.

For ^18^F-florzolotau, harmonization achieved similar threshold alignment: PET-CT optimal threshold of 1.2 SUVR versus 1.3 for harmonized PET-MRI (67% reduction from unharmonized threshold of 1.5), with excellent correlation (*r* = 1.00, slope = 0.80; Fig.6c-d). Extension to dopaminergic imaging (^18^F-FP-CIT, Parkinson’s disease patients) demonstrated quantitative equivalence in striatal binding ratios (posterior putamen: *P* = 0.864; anterior putamen: *P* = 0.313), with bias reduced from 20–26% to <7% (Extended Data Fig. 6). These results demonstrate the framework’s broad applicability across molecular targets and disease contexts, while establishing a practical foundation for consistent cross-platform diagnosis and reliable longitudinal monitoring.

## Discussion

Here, we present a physics-guided, anatomy-anchored framework that harmonizes PET-MRI quantification to PET-CT across tracers and vendors. Across 70 same-day paired scans and a 420-case multicentre cohort, the method reduced cross-platform bias by >80%, narrowing amyloid CL discrepancies from 23.6 to 4.1 (approaching the intrinsic measurement variability of PET-CT^9^) and aligning diagnostic cutoffs (amyloid: 22.6 vs. 26.7 CL; tau SUVR: 1.2 vs. 1.3). It achieved regional statistical equivalence, preserved network topology, and generalized zero-shot to ^18^F-florbetapir and ^18^F-FP-CIT without retraining. Together, this approach enables platform-agnostic thresholds and stable longitudinal monitoring when patients transition between modalities, advancing radiation-sparing quantitative PET for trials and routine care.

Our framework advances cross-platform harmonization through three architectural innovations. First, ViT-based Autoencoder encoding captures long-range inter-regional dependencies essential for attenuation modeling, overcoming locality constraints of convolutional networks^30,31,44^. Second, contrastive learning aligns MRI-derived features to the CT attenuation domain while suppressing spurious correlations^45,46^, bridging the modality gap where CT directly measures tissue density whereas MRI estimates it indirectly^18,23^. Third, anatomically guided residual correction emphasizes generalizable imaging physics over tracer-specific patterns, enabling zero-shot performance on unseen tracers (^18^F-florbetapir, ^18^F-FP-CIT)^24,47^. These design choices enable tracer-agnostic harmonization while preserving biological signal topology.

These technical advances translate into three clinically relevant outcomes. First, harmonization preserves inter-regional correlation structure (network-level *r* ≥ 0.95; Fig. 3d-4d), maintaining spatial patterns critical for Braak staging and differential diagnosis. The resulting precision exceeds thresholds for detecting annual progression (amyloid: ∼7 CL/year^48^; tau: ∼0.05 SUVR/year^49^). Second, cross-platform bias is reduced from 10–25% (typical in practice^15–17^), enabling direct application of PET-CT-derived diagnostic cutoffs^10–12,43^. Third, the tracer-agnostic architecture generalizes to emerging radiotracers targeting synaptic density, neuroinflammation, and microglial activation^50^, establishing harmonization as shared infrastructure for multitracre profiling.

**Fig. 4.**
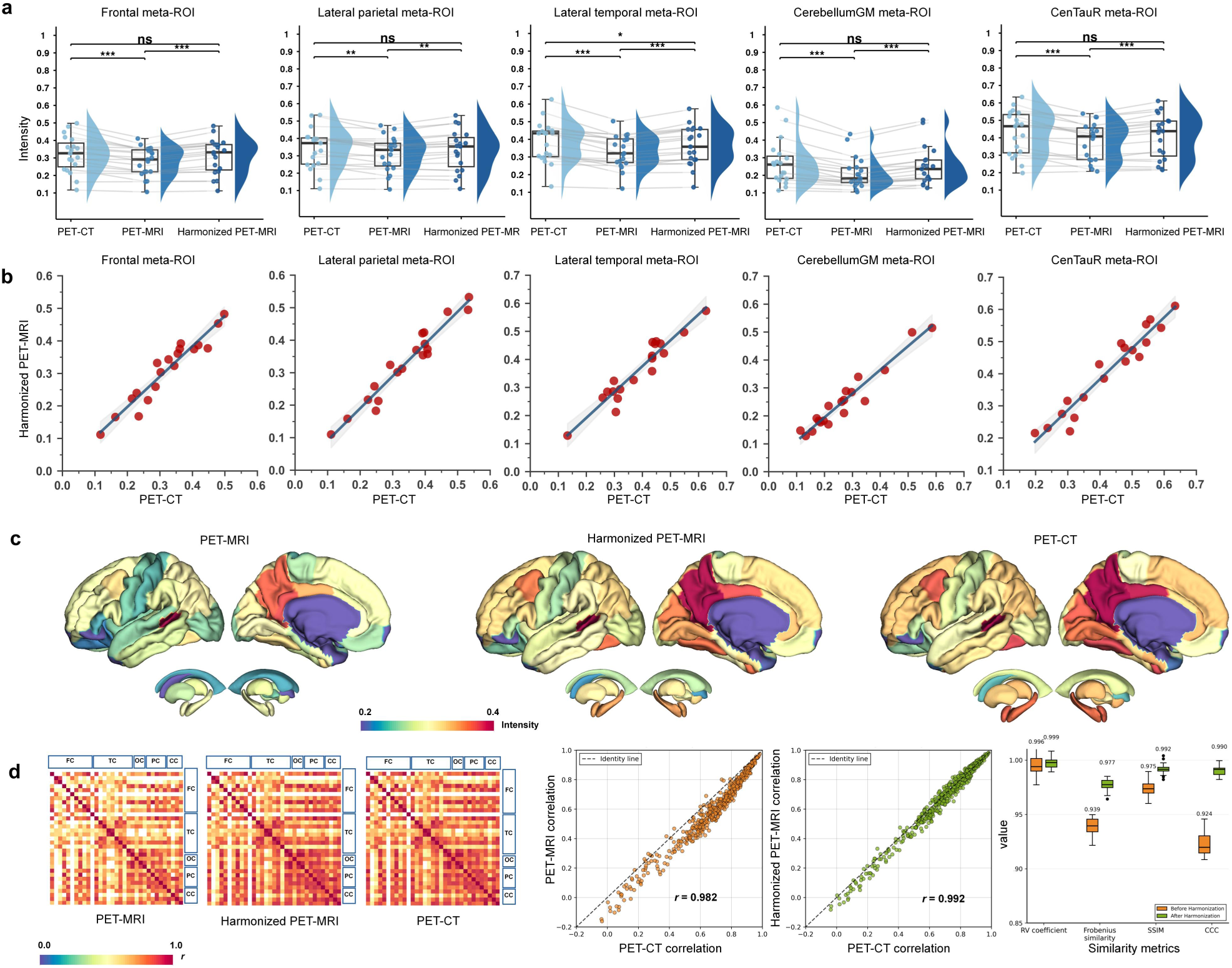
Cross-platform harmonization of ^18^F-florzolotau quantification. **a**, Regional normalized intensity distributions comparing PET-CT (reference), unharmonized PET-MRI, and harmonized PET-MRI across five brain regions. Violin plots: median (center line) and quartiles. Statistical comparisons: Wilcoxon signed-rank test with Bonferroni correction. \*\*\**P* < 0.001; \*\**P* < 0.01; \**P* < 0.05; *ns* not significant (*P* > 0.05). Harmonization achieved statistical equivalence with PET-CT across all regions. **b**, Cross-platform concordance for five regions and CenTauR meta-ROI. Each panel: scatter plot (harmonized PET-MRI versus PET-CT), regression line, Pearson correlation coefficient (*r*), coefficient of determination (*R*^2^), and *P* value. Exceptional concordance observed for CenTauR meta-ROI (*r* = 0.96, slope = 0.96) with near-unity slopes (0.85–0.99) across regions, confirming preserved spatial distribution patterns essential for Braak staging. All *P* < 0.001. See Extended Data Table 5 for detailed parameters. **c**, Cortical surface projections showing preserved tau distribution patterns. Color scale: normalized intensity (0.2–0.4). **d**, Inter-regional correlation matrices (33 regions). Left: correlation matrices for unharmonized and harmonized PET-MRI versus PET-CT. Middle: matrix-level correlation: *r* = 0.98 to 0.99 (*P* < 0.001). Region-pair analysis (Extended Data Fig. 5b) shows 91% (30/33) of regions with improved concordance.

**Fig. 5.**
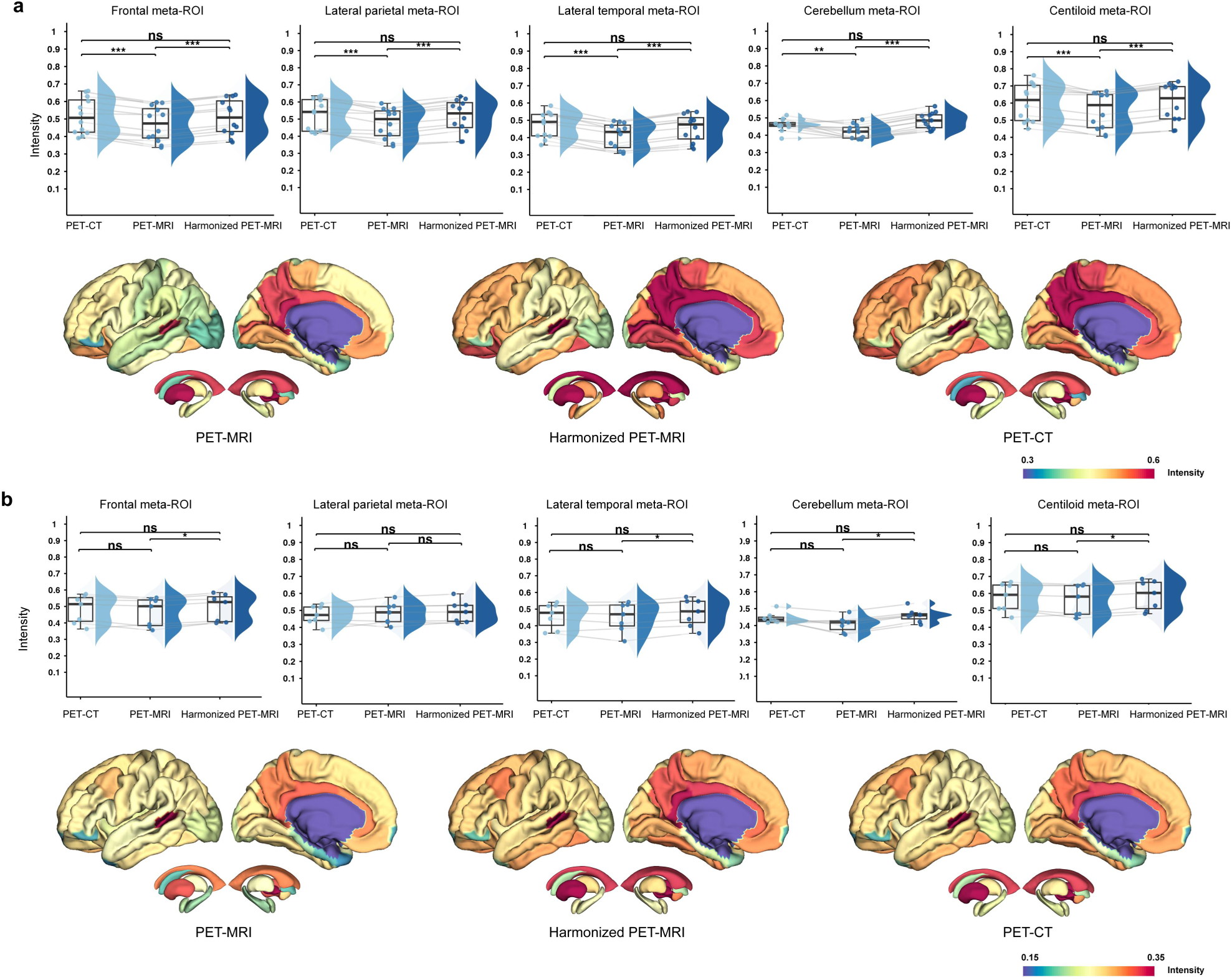
Zero-shot generalization to ^18^F-florbetapir across independent sites and vendors. Framework tested on a held-out tracer at two independent centers without retraining. a, Site 2 (Siemens Biograph mMR, *N* = 12). Top: regional normalized intensity distributions for unharmonized PET-MRI, harmonized PET-MRI, and PET-CT reference across five composite regions. Violin plots: median (center line) and quartiles. Unharmonized PET-MRI showed significant platform bias (all *P* < 0.05, Wilcoxon signed-rank test), which harmonization eliminated (all *P* > 0.05). Bottom: cortical surface projections showing preserved amyloid distribution patterns. b, Site 1 (United Imaging uMI 780, *N* = 7). Top: regional normalized intensity distributions for unharmonized PET-MRI, harmonized PET-MRI, and PET-CT reference across five composite regions. Violin plots: median (center line) and quartiles. Unharmonized PET-MRI showed significant platform bias (all *P* < 0.05, Wilcoxon signed-rank test), which harmonization eliminated (all *P* > 0.05). Bottom: cortical surface projections showing preserved amyloid distribution patterns. Consistent performance across vendors confirms physics-based generalization rather than tracer-specific overfitting. Color scale: normalized intensity.

**Fig. 6.**
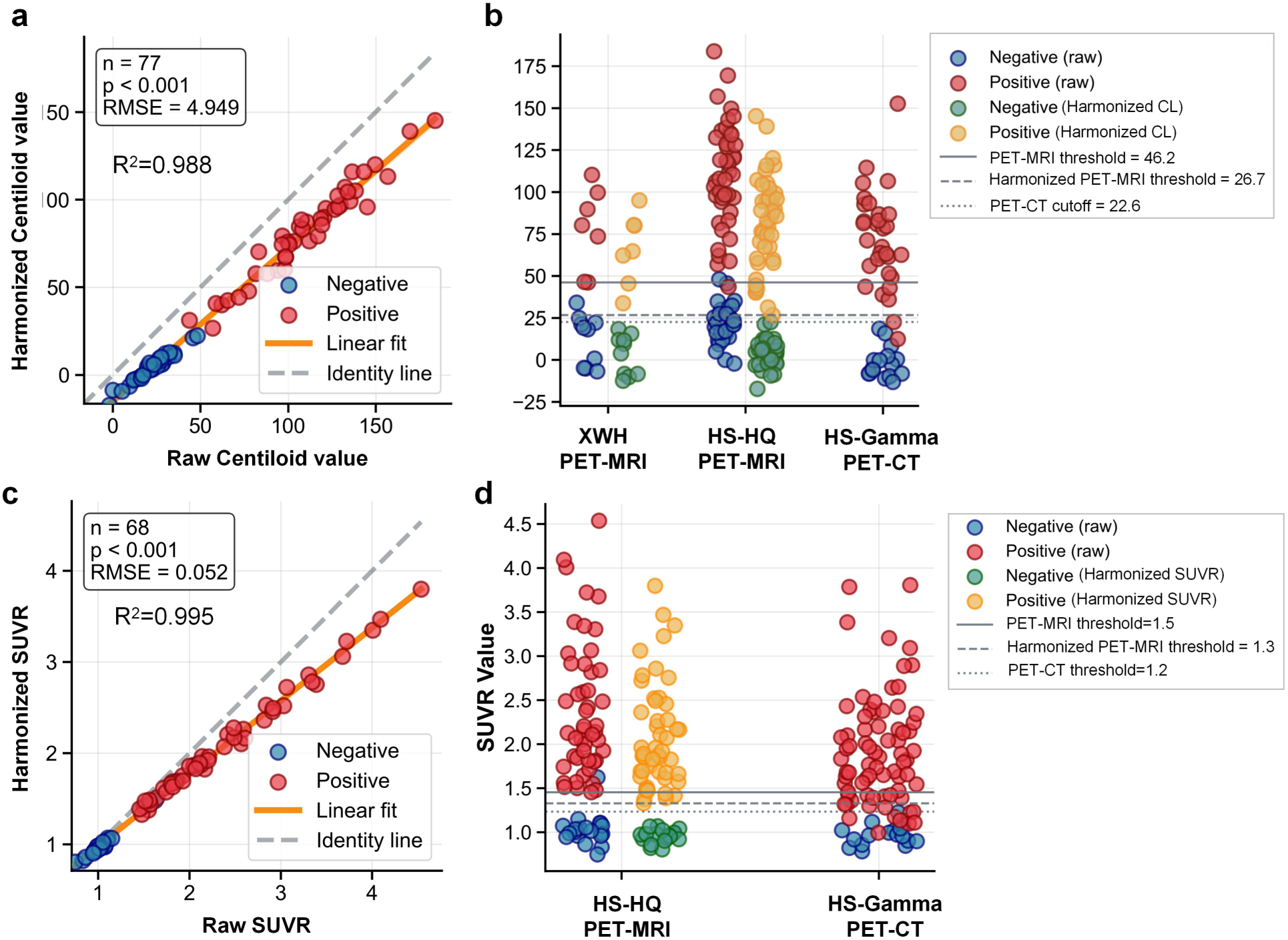
Multicentre validation of clinical threshold alignment. **a**, Centiloid distribution for ^18^F-florbetaben across PET-CT, unharmonized PET-MRI, and harmonized PET-MRI (*N* = 420, three sites, four scanners). Box plots: median (center line), IQR (box), 1.5× IQR (whiskers). Dashed lines: optimal diagnostic thresholds (PET-CT: 22.6; unharmonized PET-MRI: 46.2; harmonized PET-MRI: 26.7). Harmonization achieved 83% bias reduction. **b**, Scatter plot of harmonized PET-MRI versus PET-CT Centiloid values. Regression line (red): *r* = 0.99, slope = 0.92, RMSE = 4.95 units. Dashed lines: clinical cutoffs. Harmonized discrepancy (4.1 units) approaches test-retest precision. **c, d**, Similar analysis for ^18^F-florzolotau SUVR thresholds. PET-CT: 1.2; harmonized PET-MRI: 1.3 (67% reduction from unharmonized: 1.5). Correlation: *r* = 1.00, slope = 0.80.

The clinical implications are immediate for AD therapy monitoring. Anti-amyloid monoclonal antibodies (for example, lecanemab, donanemab) require reliable longitudinal PET to guide treatment decisions^4,5^. In the Clarity AD trial, lecanemab yielded -59 CL over 18 months (treatment: -55 CL; placebo: +4 CL)^4^. Unharmonized cross-platform bias of this magnitude approaches the net treatment effect, potentially confounding response assessment when patients transition between scanners. Our framework addresses this challenge by reducing platform discrepancy to a level approaching test-retest variability^9^, thereby enabling quantitative interchangeability between PET-MRI and PET-CT for treatment monitoring. This achievement has three practical implications. First, unified diagnostic thresholds eliminate scanner-dependent misclassification of borderline cases, where small differences near cutoffs (for example, 25–35 CL units) determine treatment eligibility^6,10,43^. Second, patients can transition between platforms during multi-year therapy without compromising longitudinal reliability, which is increasingly relevant as institutions adopt PET-MRI for radiation-sensitive repeat scanning. Third, multicentre trials can integrate PET-MRI sites previously excluded due to quantification uncertainties, expanding enrollment and reducing radiation burden in large-scale studies.

Existing harmonization strategies, including EARL accreditation and vendor-specific calibrations, improve cross-site consistency but remain limited by tracer-specific tuning and manual intervention. ComBat-based statistical harmonization^51^ and physics-driven approaches^52^ have shown promise but do not generalize across tracers without retraining. Our framework’s zero-shot performance on held-out tracers distinguishes it from prior methods, demonstrating that learning CT-anchored anatomical representations enables tracer-agnostic correction. Multicentre validation across four vendors without site-specific recalibration^53,54^ further supports scalable deployment.

Several limitations warrant consideration. Our clinical validation is retrospective and focused on neurodegenerative cohorts; generalizability to other indications (oncology, cardiology) and to pediatric populations remains to be established. Although testing spanned four vendors, emerging platforms such as total-body PET-MRI were not evaluated. Zero-shot generalization was shown for^18^F-florbetapir and ^18^F-FP-CIT, whereas tracers with markedly different kinetics (e.g., ^11^C-labeled compounds) require dedicated validation. Prospective studies should assess whether harmonization reduces boundary misclassification, stabilizes longitudinal trajectories during platform transitions, and improves trial efficiency (smaller samples or shorter follow-up). Integration with foundation-model pretraining may further enhance cross-scanner robustness and enable low-data adaptation to new tracers, and the physics-driven design may extend to cardiac and whole-body PET-MRI with modest domain-specific fine-tuning^55^. Broader deployment will also require validation across diverse clinical workflows and integration into vendor-neutral reconstruction pipelines.

In summary, this framework establishes quantitative equivalence between PET-MRI and PET-CT across multiple tracers and vendors while preserving biological signal topology. By enabling direct threshold transfer and reliable longitudinal monitoring, the approach transforms PET-MRI from a platform requiring tracer-specific validation into a quantitatively interchangeable modality. This advances radiation-sparing imaging workflows for vulnerable populations, multicentre trial integration, and longitudinal treatment monitoring, establishing harmonized PET-MRI as practical infrastructure for molecular imaging-guided precision medicine^7,14,56^.

## Methods

### Ethical approval and study design

This study was approved by the Institutional Review Board of Huashan Hospital, Fudan University (approval no. KY2024-738) and conducted in accordance with the Declaration of Helsinki. We designed a three-phase investigation: (1) prospective paired-acquisition model development, (2) prospective external validation with unseen tracer, and (3) retrospective multicentre clinical evaluation across four sites with multiple scanner platforms (Table 1). Written informed consent was obtained from prospective participants; retrospective data use was approved under institutional guidelines for de-identified data.

### Participants and data acquisition

#### Development cohort

70 patients underwent paired same-day PET-CT and PET-MRI at Huashan Hospital, Hongqiao Campus (*site 1*, *HS-HQ*) between April 2024 and April 2025: ¹⁸F-FDG (*N* = 20) for metabolic imaging, ¹⁸F-florbetaben (*N* = 20) for amyloid imaging, and ¹⁸F-florzolotau (*N* = 30) for tau imaging.

#### External validation cohort

Nineteen participants with paired ¹⁸F-florbetapir PET-CT and PET-MRI acquired at two independent centers: HS-HQ (*site 1*, *N* = 7) and Anhui Provincial Hospital (AH-PH, *site 2*, *N* = 12).

#### Clinical evaluation cohort

420 participants from three sites (*sites 1, 3-4*) with diverse scanner configurations: (1) ¹⁸F-florbetaben PET-MRI and PET-CT imaging (*N* = 143); (2) ¹⁸F-florzolotau PET-MRI and PET-CT imaging (*N* = 160); and (3) ¹⁸F-FP-CIT PET-MRI and PET-CT imaging (*N* = 117) for dopaminergic extension assessment.

#### Image acquisition protocol

To minimize biological variability, participants underwent same-day sequential PET-CT and PET-MRI acquisitions using a single radiotracer injection. The imaging sequence was randomized, with inter-scan intervals maintained at <5-7 minutes to ensure negligible changes in tracer distribution (Fig. 1). Imaging was performed at tracer-specific optimal timepoints: 70 minutes post-injection for ^18^F-florbetaben, ^18^F-florzolotau, and ^18^F-FP-CIT; 30 min for ^18^F-FDG and ^18^F-florbetapir. All acquisitions employed 20-min scan durations to ensure adequate count statistics while maintaining patient comfort (Supplementary materials).

### Image preprocessing and quality control

All imaging data underwent standardized preprocessing using Statistical Parametric Mapping (SPM12, Wellcome Trust Centre for Neuroimaging) in MATLAB R2022a. PET images were rigidly co-registered to corresponding T1-weighted MR images using normalized mutual information. T1 images underwent unified segmentation and spatial normalization to Montreal Neurological Institute (MNI) space, with resulting deformation fields applied to co-registered PET images^57,58^. Normalized PET images were smoothed with a 4mm FWHM Gaussian kernel. Quality control included visual inspection to verify anatomical correspondence. For model input, intensity values were normalized to [0, 1] range, serialized into 2D axial slices, and padded to a size of 128 × 128 pixels.

### PET harmonization framework

#### Architecture overview

We developed a unified harmonization framework to address quantification discrepancies between PET-MRI and PET-CT across multiple tracers, eliminating the need for tracer-specific models (Fig. 1). The framework employs a three-stage progressive training strategy that leverages anatomical information from CT images to guide PET-MRI harmonization.

#### Stage 1: CT anatomical feature learning

The first stage learns CT feature representations directly associated with PET imaging to provide anatomical prior knowledge for the subsequent stage. A ViT-based autoencoder architecture^32^ is employed to capture both local anatomical details^59^ and global structural context essential for accurate PET quantification imaging. CT images are partitioned into non-overlapping 4 × 4 patches, which are subsequently processed by the ViT encoder *E_CT_* to generate latent feature representations. The encoder and decoder each consist of 8 transformer layers. The encoder transforms each CT image into a feature matrix *F_CT_* ∈ R*^M^*^×*D*^ , where *M =* 1024 represents the number of patches and *D = 512* denotes the embedding dimension. The ViT architecture employed multi-head self-attention mechanisms (with 8 heads per layer) to capture long-range dependencies within anatomical structures, enabling understanding of complex spatial relationships across brain regions. Position embeddings preserved spatial information during patch-based processing, ensuring anatomical locality maintenance. The reconstruction objective was formulated as:

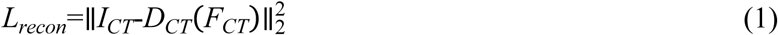

where *I_CT_* represents the input CT image and *D_CT_* denotes the decoder network.

#### Stage 2: Cross-modal feature alignment

Given that CT images provide comprehensive tissue density information essential for PET attenuation correction, MR images are inherently limited in this capacity, leading to systematic quantification errors in PET imaging^23^. To address this limitation, the second stage implements anatomical knowledge transfer from CT to MR images through a novel patch-level feature alignment framework, establishing a consistent imaging foundation for subsequent PET quantification harmonization. Specifically, an MR encoder *E_MR_* with identical architecture to the CT encoder from stage one is trained to learn MR feature representation *F_MR_* that align with the pre-trained CT feature space, thereby enabling accurate PET imaging knowledge acquisition. The alignment is achieved through a novel dual-component contrastive loss combining positive and negative pair constraints. The positive pair loss encourages feature correspondence between spatially corresponding regions by minimizing the distance between CT and MR patch representations that capture the same anatomical structures, thereby ensuring that the learned MR features preserve the spatial-anatomical relationships inherent in the CT domain, calculated as:

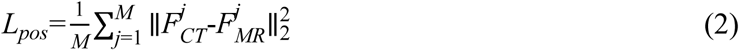

where *F_CT_^j^* and *F_MR_^j^* represent the *j*-th patch features from CT and MR images, respectively.

The negative pair loss prevents erroneous alignments between non-corresponding regions by maximizing the distance between anatomically distinct patch pairs, which is crucial for maintaining feature specificity and preventing the model from learning spurious correlations that could compromise the fidelity of cross-modal knowledge transfer:

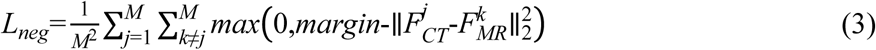

The complete contrastive loss combines both components to ensure robust feature alignment across the entire image domain:

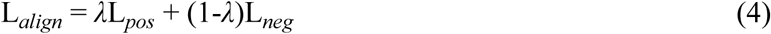

where *λ* balances the contribution of positive and negative pair constraints.

#### Stage 3: PET quantification harmonization

The final stage integrates the aligned MR features into a comprehensive harmonization framework designed to transform PET-MR images to achieve PET-CT-equivalent quantification. The architecture employs a U-Net backbone (Extended Data Fig. 7 for the detailed network architecture) enhanced with anatomical guidance to leverage the cross-modal feature alignment established in the previous stage.

The input PET-MR image is first processed through an encoder comprising Convolutional Block Attention Module (CBAM) blocks^60^ to generate multi-scale latent representations *F_ι_*. These encoder blocks progressively extract hierarchical features while maintaining spatial information through convolution and attention mechanisms.

An anatomical-guided block subsequently refines these representations by incorporating the aligned MR features *F_MR_* obtained from the second stage. This integration operation combines the PET-MR features with imaging priors through a feature fusion mechanism:

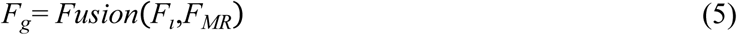

where *F_g_* represents the anatomically-informed features that preserve both functional PET information and structural anatomical context. The fusion operation employs cross-attention mechanisms to dynamically integrate multi-modal features, where PET-MR latent representations *F_ι_* serve as queries that attend to aligned MR features *F_MR_* functioning as keys and values, enabling adaptive feature weighting based on local anatomical context.

The decoder reconstructs high-resolution feature maps through symmetric CBAM blocks and skip connections, preserving fine-grained details essential for accurate quantification. The skip connections facilitate direct information flow between corresponding encoder and decoder layers, ensuring that high-frequency details are preserved during the reconstruction process. Each decoder block applies up sampling followed by feature refinement through attention mechanisms, gradually recovering spatial resolution while maintaining semantic consistency. The final harmonized image is obtained through residual learning:

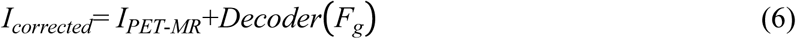

where *I_PET_*_-*MR*_ represents the input PET-MR image and *Decoder*(*F_g_*) denotes the harmonization residual generated by the decoder network. This residual formulation ensures that the framework learns to harmonize quantification discrepancies while maintaining the inherent tracer distribution patterns.

The complete loss function combines pixel-wise reconstruction accuracy with perceptual similarity:

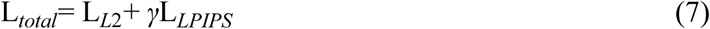

where L*_L_*_2_ represents the pixel-wise *l*_2_ loss between corrected and reference PET-CT images, L*_LPIPS_*denotes the learned perceptual image patch similarity loss, and *γ* balances the contribution of perceptual and pixel-wise objectives. This composite loss ensures both quantitative accuracy and perceptual quality of the corrected images.

#### Implementation and training details

The framework is implemented using PyTorch and trained on NVIDIA A100 GPUs with mixed precision to enhance computational efficiency. The margin parameter in the negative pair loss is empirically set to 1.0, and the balancing parameter *λ* is set to 0.5. The perceptual loss weight *γ* is set to 0.1 to balance pixel-wise and perceptual objectives.

To ensure a comprehensive evaluation, all experiments are conducted using a five-fold cross-validation at the subject level (four ^18^F-FDG, fou ^18^F-Florbetaben, and six ^18^F-Florzolotau imaging per fold). Training is conducted using the Adam optimizer and Cosine Annealing learning rate scheduling. The framework undergoes sequential training across the three stages, with each stage optimized for convergence before proceeding to the next.

The detailed training hyperparameters for each stage were as follows: Stage 1 was trained for 200 epochs with a batch size of 8 for CT anatomical feature learning, using an initial learning rate of 1 × 10^-4^ that was gradually decreased to 1 × 10^-6^ after 20 warm-up epochs. Stage 2, focused on cross-modal feature alignment, was trained for 100 epochs with a batch size of 8, employing an initial learning rate of 1 × 10^-3^ that decayed to 1 × 10^-4^ after 10 warm-up epochs. Finally, Stage 3 required 200 epochs with a batch size of 32 for the final PET quantification harmonization network training, starting from a learning rate of 1 × 10^-4^ and decaying to 1 × 10^-5^ after 10 warm-up epochs.

### Quantitative evaluation

#### Image quality assessment

Framework performance was evaluated using peak PSNR and structural SSIM^61^ computed between harmonized PET-MRI and reference PET-CT images using:

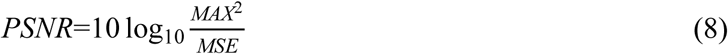

where *MAX* represent the maximum possible pixel value and *MSE* denotes the mean squared error between the harmonized and ground-truth images. The SSIM that captures luminance, contrast, and structural information was computed using:

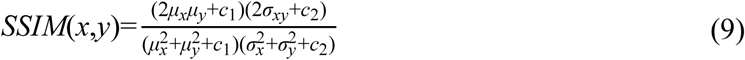

where *μ_x_* and *μ_y_* are the mean intensities of harmonized and ground-truth images, *σ*^2^ and *σ*^2^ are their variances, *σ_xy_* is the covariance, and *c*_1_ , *c*_2_ are stabilization constants. Higher PSNR values indicate better pixel-wise fidelity, while SSIM values closer to 1.0 represent superior structural preservation between harmonized PET-MR and real PET-CT images, thereby demonstrating higher harmonization efficacy of our framework.

Benchmarking compared our framework against five established methods: unprocessed PET-MRI (baseline), SwinIR^39^, Restormer^40^, DRMC^41^, and AIMIR^42^.

#### Regional quantification

Regional analysis employed FreeSurfer-derived anatomical parcellations: Desikan-Killiany atlas (68cortical regions)^62^, ASEG atlas (14 subcortical structures)^63^, Tian atlas (anterior and posterior putamen)^64^, and composite meta-ROIs for large-scale cortical regions. Two complementary quantification metrics were computed for all regions: (1) Normalized intensity measurements (mean value averaged across voxels without reference normalization), and (2) SUV ratios (SUVR, regional SUV to tracer-specific reference regions: whole cerebellum for amyloid tracers, cerebellar gray matter for tau tracers, and pons for FDG)^10,14^, providing relative quantification that accounts for inter-subject differences in tracer delivery and clearance. Standardized quantification scales included CL value for amyloid PET^56^ and CenTauR for tau PET^13^ calculated using standard equation incorporating whole cerebellum reference and cortical target regions. CenTauR values utilized temporal meta-ROI uptake normalized to cerebellar gray matter.

All analyses were performed in parallel using both normalized intensity and SUVR to validate robustness across quantification frameworks. This dual-metric approach confirmed that harmonization effects were independent of the specific quantification method employed.

### Statistical analysis

Quantitative agreement was evaluated through comprehensive three-way comparisons (PET-CT reference, unharmonized PET-MRI, harmonized PET-MRI) to quantify both baseline platform discrepancies and harmonization improvements.

#### Regional comparisons

Pairwise statistical testing employed Wilcoxon signed-rank tests^65,66^ with Bonferroni correction (significance threshold *P* < 0.05). Three contrasts were evaluated for each brain region and tracer: (1) unharmonized PET-MRI versus PET-CT (baseline differences), (2) harmonized PET-MRI versus PET-CT (residual differences), and (3) unharmonized versus harmonized PET-MRI (direct harmonization effects).

#### Cross-platform calibration

Linear regression quantified relationships between harmonized PET-MRI and PET-CT measurements, reporting Pearson correlation coefficients (*r*), coefficients of determination (*R*^2^), regression slopes, and intercepts. Ideal harmonization was defined as near-unity slopes (0.9-1.1) with high correlations (*r* > 0.90), indicating eliminated systematic bias while preserving inter-subject variability.

#### Measurement agreement

Bland-Altman analyses assessed systematic bias (mean difference) and measurement variability (95% LoA, LoA = mean ± 1.96×SD) for the three pairwise comparisons above. Analyses were performed as pooled assessments combining measurements across tracer-specific composite regions and subjects. Successful harmonization was defined by: (1) substantial mean bias reduction, ideally to <5% of baseline; (2) transformation from asymmetric baseline LoA to symmetric post-harmonization distributions centered near zero; and (3) maintained or improved measurement precision.

#### Inter-regional correlation preservation

For each modality, correlation matrices representing inter-regional relationships were constructed. Network topology similarity was quantified using matrix-level correlation, RV coefficient, Frobenius similarity, SSIM, and concordance correlation coefficient. Mantel tests (10,000 permutations) assessed statistical significance of matrix correlations.

#### Clinical validation

Optimal thresholds were determined using Youden’s index. Threshold shifts <5% from PET-CT reference were considered clinically acceptable. For ^18^F-FP-CIT dopaminergic imaging, striatal binding ratios were compared across modalities to validate preservation of clinically relevant features. All analyses were performed using R v4.3.0 and Python 3.9 with NumPy, SciPy, and scikit-learn. Statistical significance was set at *P* < 0.05 for two-sided tests.

##### Acknowledgements

This study was supported by the National Natural Science Foundation of China (grant no. 82394434,82272039, and 82021002) and STI2030-Major Projects (grant no. 2022ZD0211606) to C.T.Z; the National Natural Science Foundation of China (grant no. 82394432) to K.S; the Shanghai Medical Innovation & Development Foundation (grant no. SMIDF-150-2025A18) to H.Z. The computation resources in this article were provided by AI4S Initiative and HPC Platform of ShanghaiTech University.

## Author contributions

J.W., A.Z., C.Z. and Q.W. participated in conceptualization, methodology, resources, writing of the original draft, supervision and funding acquisition. J.W., A.Z., and H.H. participated in the conceptualization, methodology, and formal data analysis. J.W., A.Z., and H.H. participated in the writing of the original draft. J.W., Y.Z. H.Z., J.L., C.L., Q.X., J.L., C.J, M.N., and Y.G. collected and organized data. H.Z., J.J., M.W., K.S., M.T., and D.S. provided critical comments and reviewed the paper. All authors contributed to the research, editing and approval of the paper.

## Competing interests

D.S. are consultant and employee of Shanghai United Imaging Intelligence Co., Ltd. The company had no role in designing or performing the surveillance, nor in analysing or interpreting the data. The other authors declare no competing interests.

**Correspondence and requests for materials** should be addressed to Chuantao Zuo and Qian Wang.

## Data availability

The main data supporting the results in this study are available within the article and its Supplementary Information. Individual-level patient data are protected because of patient privacy; they are accessible with the consent of the data management committee from institutions and are not publicly available. Requests for the non-profit use of the images and related clinical information should be sent to C.T.Z (zuochuantao@fudan. edu.cn). The data management committee will then review all the requests and grant permission (if successful). A formal data transfer agreement will be required upon approval. All data shared will be deidentified. Source data are provided with this paper.

## Code availability

Our code will be released upon acceptance.

---

**Extended Data Fig. 1.**
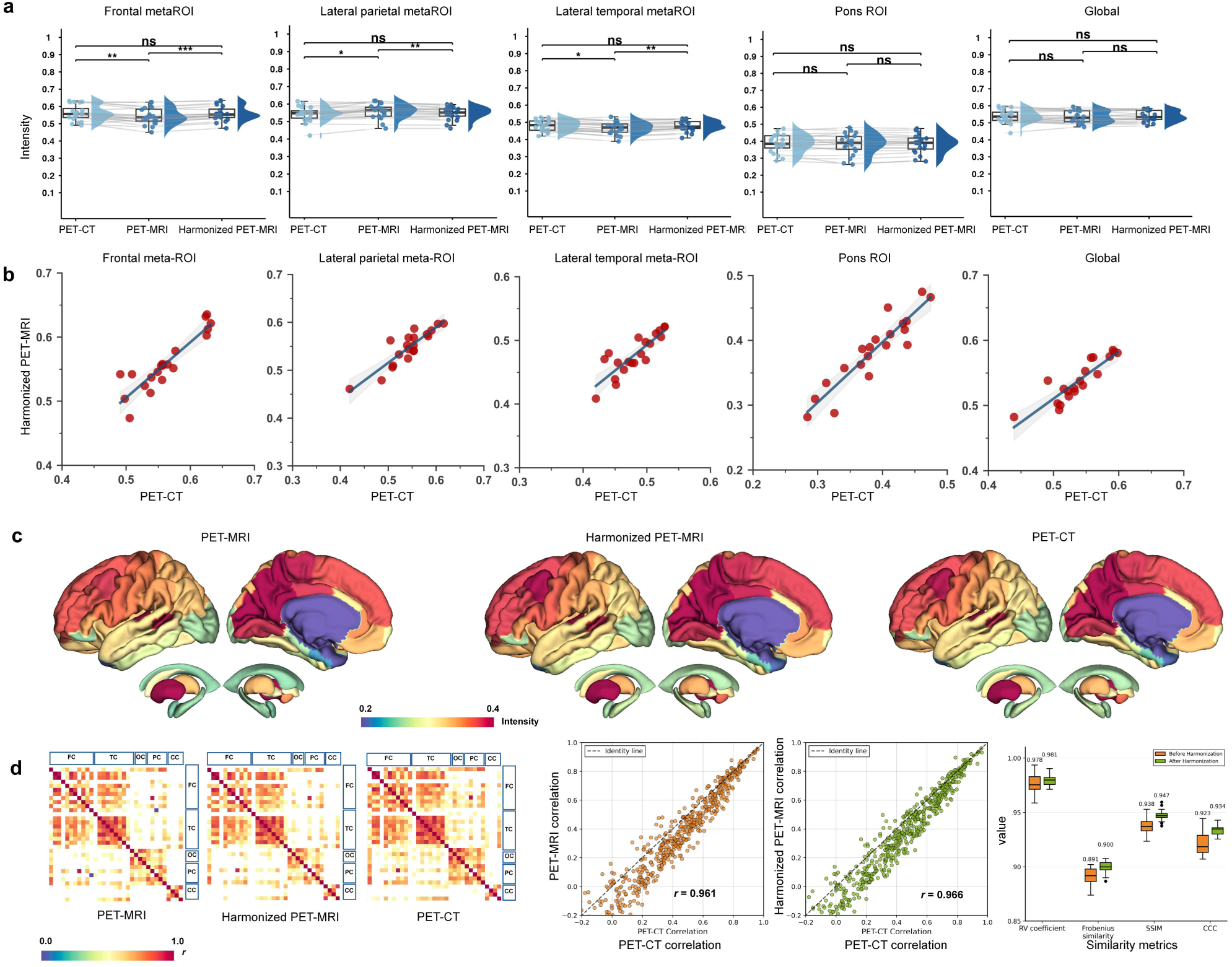
^18^F-FDG quantification harmonization. **a.** Regional normalized intensity across five brain regions comparing PET-CT reference, unharmonized PET-MRI, and harmonized PET-MRI. Violin plots show distribution density with median and quartiles. Statistical comparisons via Wilcoxon signed-rank test with Bonferroni correction: \*\*\**P* < 0.001, \*\**P* < 0.01, \**P* < 0.05, ns: *P* > 0.05. Harmonization achieved statistical equivalence with PET-CT across all regions. **b**, Cross-platform concordance analysis showing preserved inter-subject variability. Each panel shows one brain region with scatter plot, regression line, Pearson correlation (*r*), coefficient of determination (*R*^2^), and significance. Strong cross-platform concordance maintained across metabolic networks (pons reference: *r* = 0.92, slope = 0.93; regional: *r* = 0.85-0.92, slopes = 0.72-0.93), with consistently high correlations (all *r* > 0.85, all *P* < 0.001) confirming preserved inter-subject ranking patterns necessary for clinical interpretation of neurodegenerative metabolic changes. **c**, Cortical surface projections showing preservation of metabolic distribution patterns. Color scale: normalized intensity (0.2-0.4). **d**, Inter-regional correlation matrices (33 regions). left: Correlation matrices for PET-CT reference, unharmonized PET-MRI, and harmonized PET-MRI. middle: Scatter plot showing improved matrix-level correlation (*r* = 0.96→0.97, *P* < 0.001). Each point represents correlation between a unique region pair. right: Network similarity metrics. Region-pair analysis showing 79% (26/33) of regions with enhanced correlation concordance in Extended Data Fig. 5c.

**Extended Data Fig. 2.**
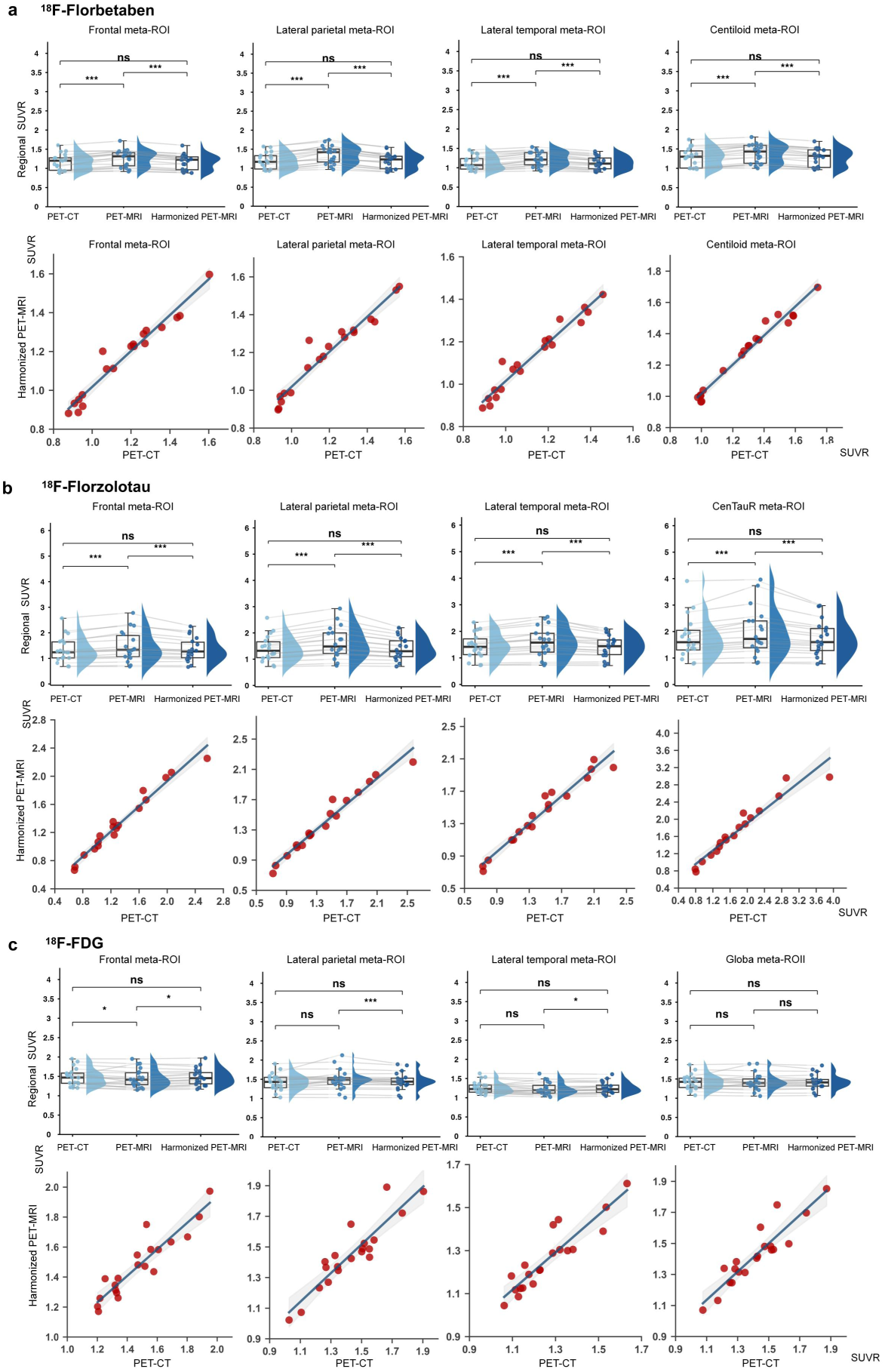
SUVR quantification demonstrates robust cross-platform harmonization. Regional SUVR harmonization performance across three PET tracers validated using tracer-specific reference regions (whole cerebellum for amyloid tracers, cerebellar gray matter for tau, pons for FDG). **a**, ^18^F-florbetaben SUVR validation. upper: Violin plots comparing SUVR distributions across five brain regions between PET-CT reference (left), unharmonized PET-MRI (middle), and harmonized PET-MRI (right). Harmonization reduced regional SUVR biases from 7.78-13.07% to -0.11-0.60%. Statistical significance: \*\*\**P* < 0.001, \*\**P* < 0.01, \**P* < 0.05, ns *P* > 0.05 (Wilcoxon signed-rank test with Bonferroni correction). lower: Linear regression analyses between harmonized PET-MRI and PET-CT SUVR values demonstrating near-perfect concordance (Centiloid mask: *r* = 0.99, slope = 0.93; regional: *r* = 0.96-0.99, slopes = 0.90-0.95; all *P* < 0.001). Detailed parameters in Extended Data Table 6. **b**, ^18^F-florzolotau SUVR validation. upper: Regional SUVR distributions showing bias reduction from 8.31-13.62% to -0.71-0.81%. lower: Linear regression demonstrating exceptional concordance (CenTauR: *r* = 0.97, slope = 0.79; regional: *r* = 0.97-0.98, slopes = 0.84-0.89; all *P* < 0.001). **c**, ^18^F-FDG SUVR validation. upper: Regional SUVR distributions with maintained low baseline biases further improved post-harmonization. lower: Strong regression agreement (*r* = 0.91-0.92, slopes = 0.87-0.95; all *P* < 0.001). Concordance between normalized intensity (Fig. 3-5) and SUVR findings confirms robust harmonization independent of quantification metric. Corresponding normalized intensity analyses in main Fig. 3-5 and Supplementary Table 2.

**Extended Data Fig. 3.**
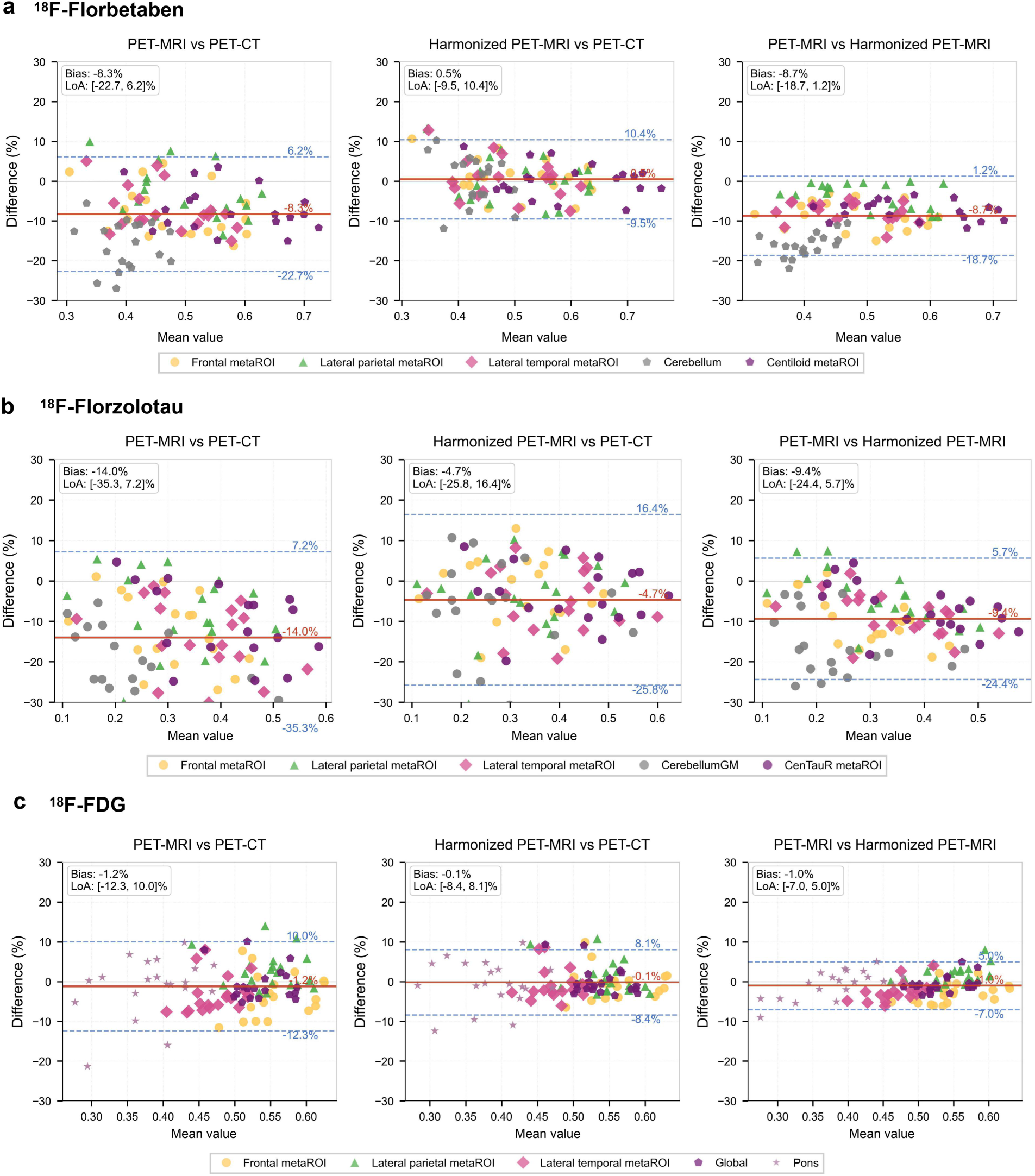
Measurement agreement analysis across platforms. Bland-Altman plots for normalized intensity comparing: unharmonized PET-MRI vs PET-CT (left), harmonized PET-MRI vs PET-CT (middle), and direct harmonization effect (right). Solid line: mean bias; dashed lines: 95% limits of agreement. Points colored by region. a, ^18^F-florbetaben: bias reduced 94% (−8.3% to 0.5%), with LoA transforming from asymmetric [-22.7%, 6.2%] to symmetric [-9.5%, 10.4%] distribution centered near zero. b, ^18^F-florzolotau: bias reduced 66% (−14.0% to -4.7%), with LoA shifting from [-35.3%, 7.2%] to [-25.8%, 16.4%]. c, ^18^F-FDG: bias reduced 92% (−1.2% to -0.1%), with LoA narrowing from [-12.3%, 10.0%] to [-8.4%, 8.1%]. Near-zero mean bias combined with reduced LoA range demonstrates enhanced agreement across both systematic and random measurement components. Region-specific values in Supplementary Table 1.

**Extended Data Fig. 4.**
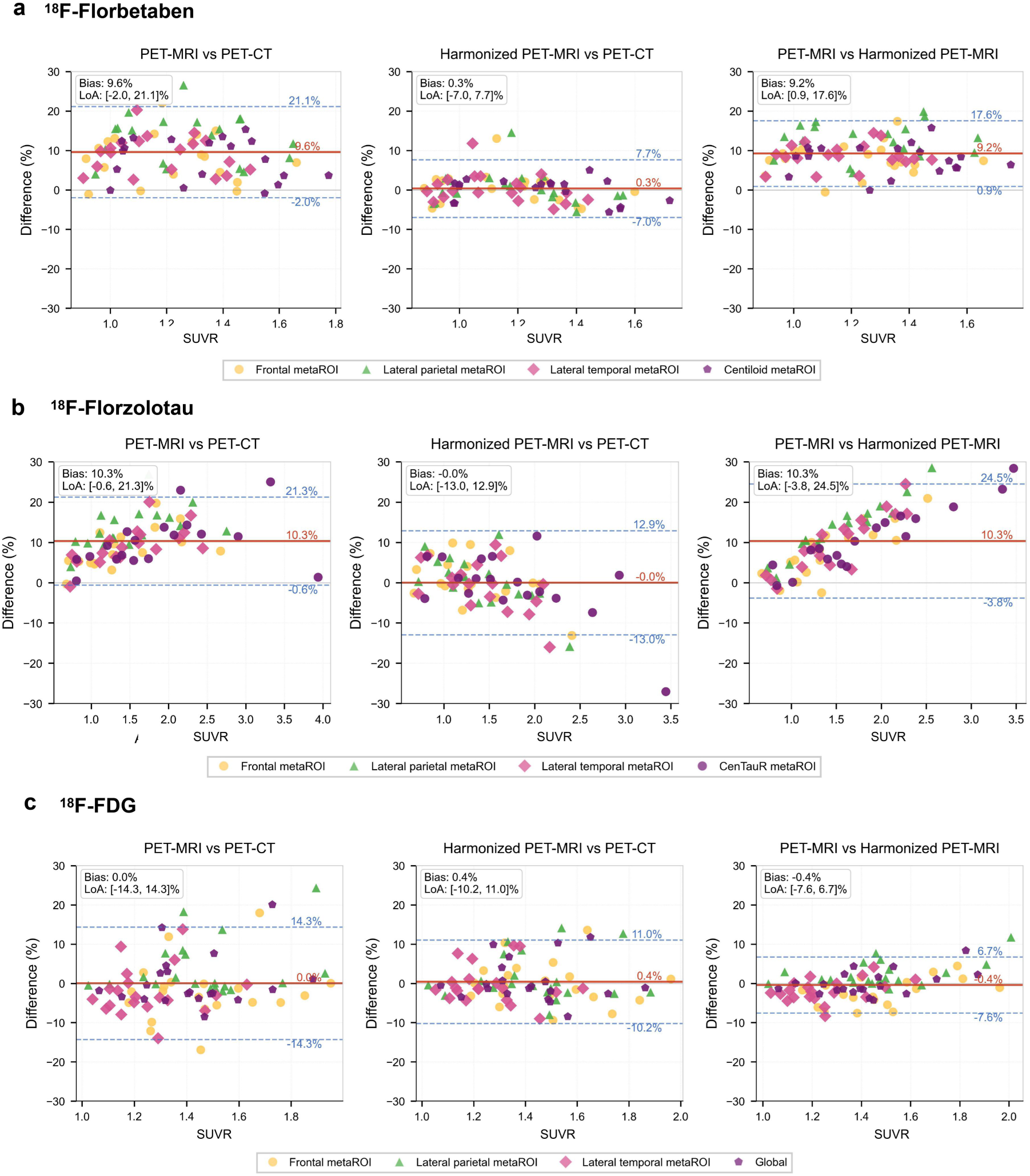
Measurement agreement analysis across platforms. Bland-Altman plots for SUVR comparing: unharmonized PET-MRI vs PET-CT (left), harmonized PET-MRI vs PET-CT (middle), and direct harmonization effect (right). Layout and statistical methods identical to main Fig. 6, with SUVR values replacing normalized intensity measurements. **a**, ^18^F-florbetaben SUVR agreement. left: Unharmonized PET-MRI vs PET-CT showing asymmetric distribution (pooled mean bias: 8.6%, LoA: ∼[-2%, 21%]). middle: Harmonized PET-MRI vs PET-CT demonstrating substantial improvement (mean bias: 0.3%, symmetric LoA: [-7%, 8%]). right: Direct comparison confirming mathematical consistency. **b**, ^18^F-florzolotau SUVR agreement. Pooled mean bias reduced from 10.3% to ∼0%, with LoA transforming from asymmetric [∼-1%, 21%] to symmetric [-13%, 13%]. **c**, ^18^F-FDG SUVR agreement. Near-zero baseline bias maintained with improved precision (LoA: [-14%, 14%] to [-10%, 11%]). Post-harmonization SUVR biases reduced to <1% for amyloid and tau imaging, with narrowed LoA across all tracers, confirming substantial improvement in cross-platform measurement agreement. Region-specific values in Supplementary Table 2.

**Extended Data Fig. 5.**
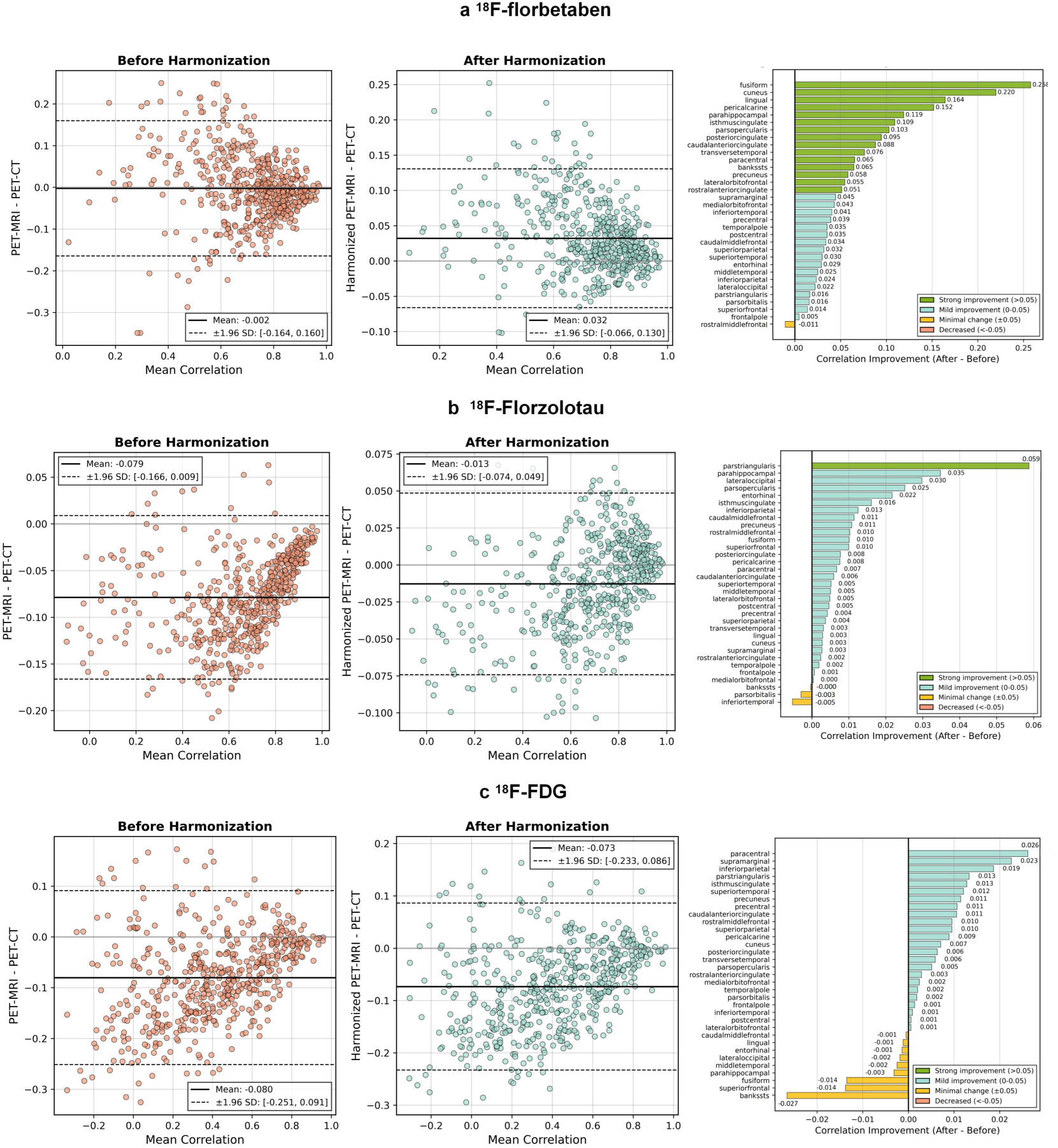
Preservation of inter-regional correlation networks. Region-pair level analysis of inter-regional correlation pattern preservation for three radiotracers. **a**, ^18^F-florbetaben correlation preservation. left: Bland-Altman plot of correlation differences between PET-CT reference and unharmonized PET-MRI, showing systematic negative bias (mean: -0.002). middle: Post-harmonization comparison demonstrating eliminated bias (mean: 0.032) with tightly centered distribution. right: Bar plot showing regional correlation improvements (Δ*r* = rharmonized - rraw) ranked by magnitude. Color coding: green (strong improvement, Δ*r* > 0.05), light teal (mild improvement, 0<Δ*r* ≤ 0.05), yellow (minimal change, |Δ*r*| ≤ 0.05), orange (decreased, Δ*r* < -0.05). 97% (32/33) of regions showed positive correlation gains, with strong improvements in cortical regions critical for amyloid distribution patterns. **b**, ^18^F-florzolotau correlation preservation. Bland-Altman analysis showed systematic bias reduction from -0.079 (unharmonized) to -0.013 (harmonized). 91% (30/33) of regions demonstrated improved correlation concordance, with strong improvements in parahippocampal, entorhinal, and temporal cortices essential for tau network topology and Braak staging. **c**, ^18^F-FDG correlation preservation. Systematic correlation bias reduced from -0.080 to -0.073, with 79% (26/33) of regions showing enhanced concordance. Strong improvements observed in paracentral, supramarginal, and inferior parietal cortices, maintaining metabolic connectivity patterns essential for neurodegenerative change interpretation. Whole-network correlation improvements detailed in main Fig. 3-4d. Preservation of inter-regional patterns confirms harmonization maintains biologically meaningful correlation structures while correcting platform-dependent quantification biases.

**Extended Data Fig. 6.**
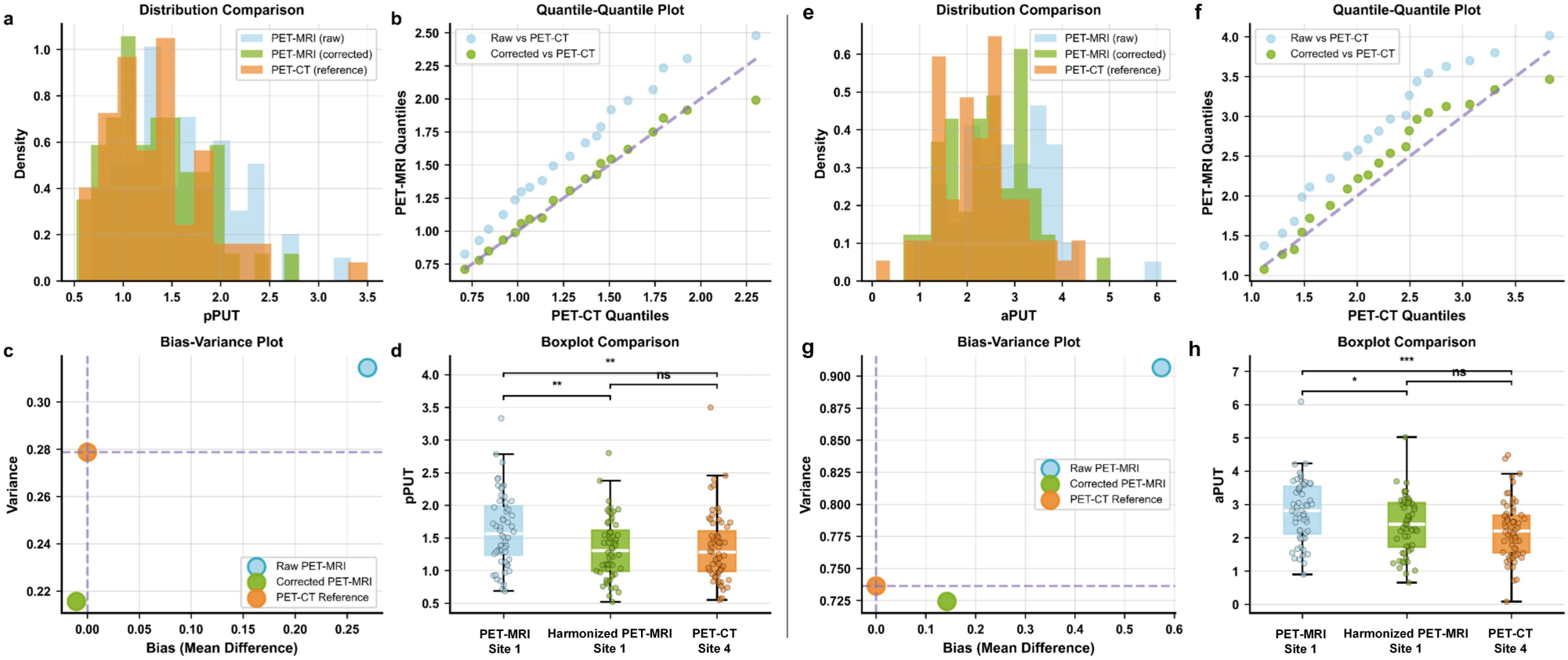
Dopaminergic imaging harmonization for movement disorders. Validation of framework applicability to ^18^F-FP-CIT dopaminergic imaging in 117 Parkinson’s disease patients, extending beyond neurodegenerative amyloid/tau/metabolic applications. **a-d**, Posterior putamen analysis. a, Distribution histograms showing convergence post-harmonization. Raw PET-MRI (blue) shows rightward shift from PET-CT reference (orange); harmonized PET-MRI (green) demonstrates restored alignment. b, Quantile-quantile plots demonstrating improved correlation (*r* = 0.996→0.998). c, Bland-Altman plot showing bias reduction from 19.98% to -0.79% with narrowed limits of agreement. d, Box plots confirming SUVR alignment: raw 1.619 ± 0.561 vs harmonized 1.339 ± 0.464 vs reference 1.350±0.528 (mean ± SD). Statistical significance: \*\*\**P* < 0.001, \*\**P* < 0.01, \**P* < 0.05, ns *P* > 0.05 (Wilcoxon signed-rank test). **e-h**, Anterior putamen analysis showing similar improvements with bias reduction from 25.64% to 6.35%. Box plots show raw 1.811 ± 0.647 vs harmonized 1.530 ± 0.508 vs reference 1.438 ± 0.530. Effect size analysis confirmed distributional alignment with Cohen’s d reduced from 0.492 to -0.021 (posterior putamen) and 0.630 to 0.165 (anterior putamen), indicating improved statistical equivalence. Kolmogorov-Smirnov tests confirmed distributional alignment post-harmonization (*P* > 0.99), enabling cross-platform diagnostic consistency for dopaminergic system assessment in movement disorder evaluation.

**Extended Data Fig. 7.**
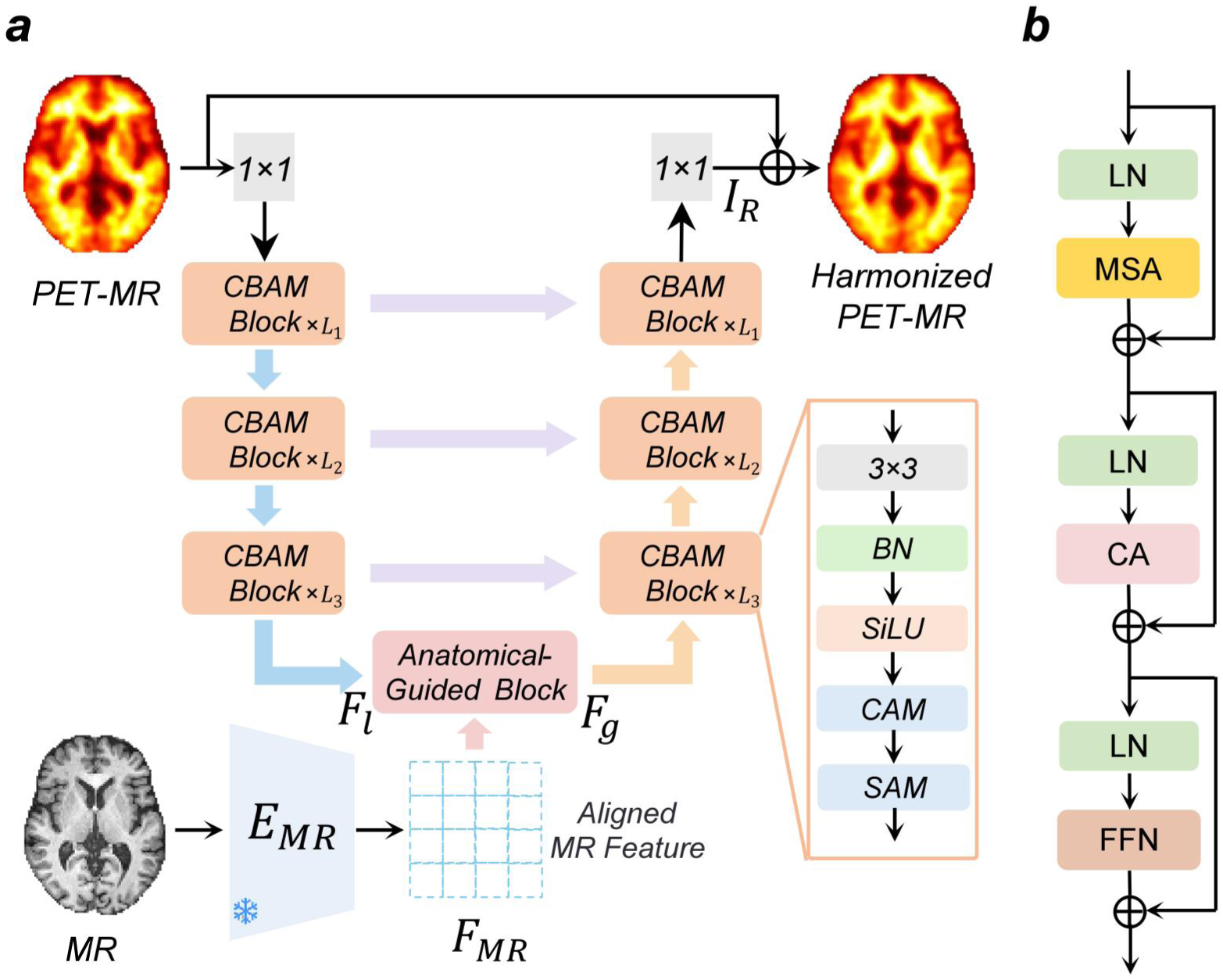
Network architecture for PET quantification harmonization. **a**, Detailed network structure for PET quantification harmonization (stage 3). The aligned anatomical features are integrated into a comprehensive harmonization network that transforms PET-MR images to achieve PET-CT-equivalent quantification while preserving tracer-specific uptake patterns across different radioligands. The architecture employs a U-Net-based backbone enhanced with anatomical guidance, comprising encoder-decoder pathways with CBAM blocks. Skip connections preserve fine-grained details during upsampling. The anatomical-guided block integrates PET features with aligned MR anatomical features obtained from the cross-modal feature alignment stage. **b**, Detailed structure of the anatomical-guided block. The block refines PET representations by incorporating aligned MR features through cross-attention mechanisms, sequentially applying layer normalization, multi-head self-attention, cross-attention, and feed-forward neural network operations with residual connections to enhance anatomical consistency in the harmonized output.

**Extended Data Table 1.**
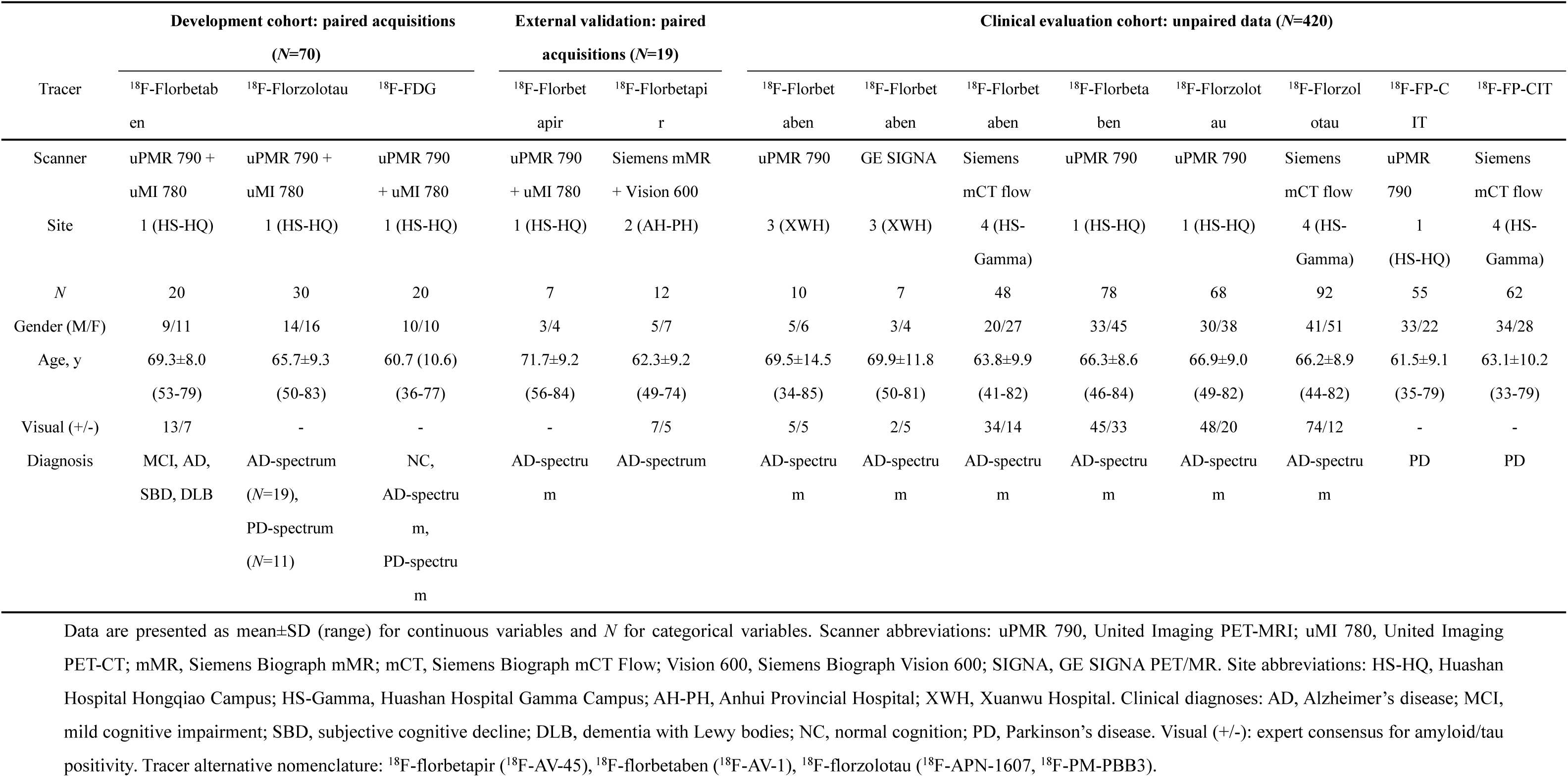
Study cohort characteristics

**Extended Data Table 2.** Quantitative performance benchmarking across radiotracers

| Tracer | Method | PSNR (dB) [range] | SSIM [range] |
| --- | --- | --- | --- |
| <sup>18</sup>F-florbetaben | PET-MR (baseline) | 31.41±1.940 [28.01-34.33] | 0.97±0.008 [0.96-0.99] |
|  | SwinIR | 33.27±1.941 [28.52-36.45] | 0.98±0.006 [0.97-0.99] |
|  | Restormer | 33.82±1.957 [27.82-37.59] | 0.98±0.006 [0.97-0.99] |
|  | DRMC | 33.42±1.775 [29.07-36.07] | 0.98±0.005 [0.97-0.99] |
|  | AIMIR | 34.07±1.942 [28.13-36.76] | 0.98±0.005 [0.97-0.99] |
|  | **Ours** | **34.80±2.133 [28.42-37.84]** | **0.98±0.005 [0.98-0.99]** |
| <sup>18</sup>F-florzolotau | PET-MR (baseline) | 34.54±4.695 [26.02-43.03] | 0.96±0.020 [0.91-0.99] |
|  | SwinIR | 36.02±3.906 [27.53-44.41] | 0.97±0.016 [0.93-0.99] |
|  | Restormer | 36.25±3.642 [29.29-43.81] | 0.97±0.014 [0.93-0.99] |
|  | DRMC | 36.45±3.922 [28.20-44.24] | 0.97±0.015 [0.93-0.99] |
|  | AIMIR | 36.36±3.828 [28.75-44.20] | 0.97±0.011 [0.94-0.99] |
|  | **Ours** | **36.77±3.670 [30.32-45.16]** | **0.98±0.011 [0.94-0.99]** |
| <sup>18</sup>F-FDG | PET-MR (baseline) | 36.18±1.582 [32.20-38.17] | 0.99±0.004 [0.98-0.99] |
|  | SwinIR | 35.81±1.712 [31.77-37.91] | 0.99±0.004 [0.98-0.99] |
|  | Restormer | 36.27±1.512 [33.03-37.72] | 0.98±0.003 [0.98-0.99] |
|  | DRMC | 36.40±1.655 [32.33-38.53] | 0.99±0.004 [0.98-0.99] |
|  | AIMIR | 36.53±1.547 [33.18-38.20] | 0.98±0.004 [0.98-0.99] |
|  | **Ours** | **37.25±1.582 [33.64-39.09]** | **0.99±0.003 [0.98-0.99]** |

PSNR and SSIM values (mean±SD [range]) comparing our framework against five established harmonization approaches across three PET tracers. Performance evaluated on held-out test set using five-fold cross-validation at subject level. Bold values indicate best performance. Our framework achieved superior performance across all tracers. PSNR, peak signal-to-noise ratio; SSIM, structural similarity index measure; PET-MR (baseline), unharmonized PET-MRI; SwinIR, Swin Transformer Image Restoration network; Restormer, Efficient Transformer for High-Resolution Image Restoration; DRMC, a network with dynamic routing for multi-center pet image synthesis; AIMIR, All-in-one medical image restoration network. Higher PSNR and SSIM values indicate better harmonization quality. Data represent mean±standard deviation with full range in brackets. The reported standard deviation reflects the sample-wise variability, computed from the metric values of all test samples aggregated across the five folds.

**Extended Data Table 3.**
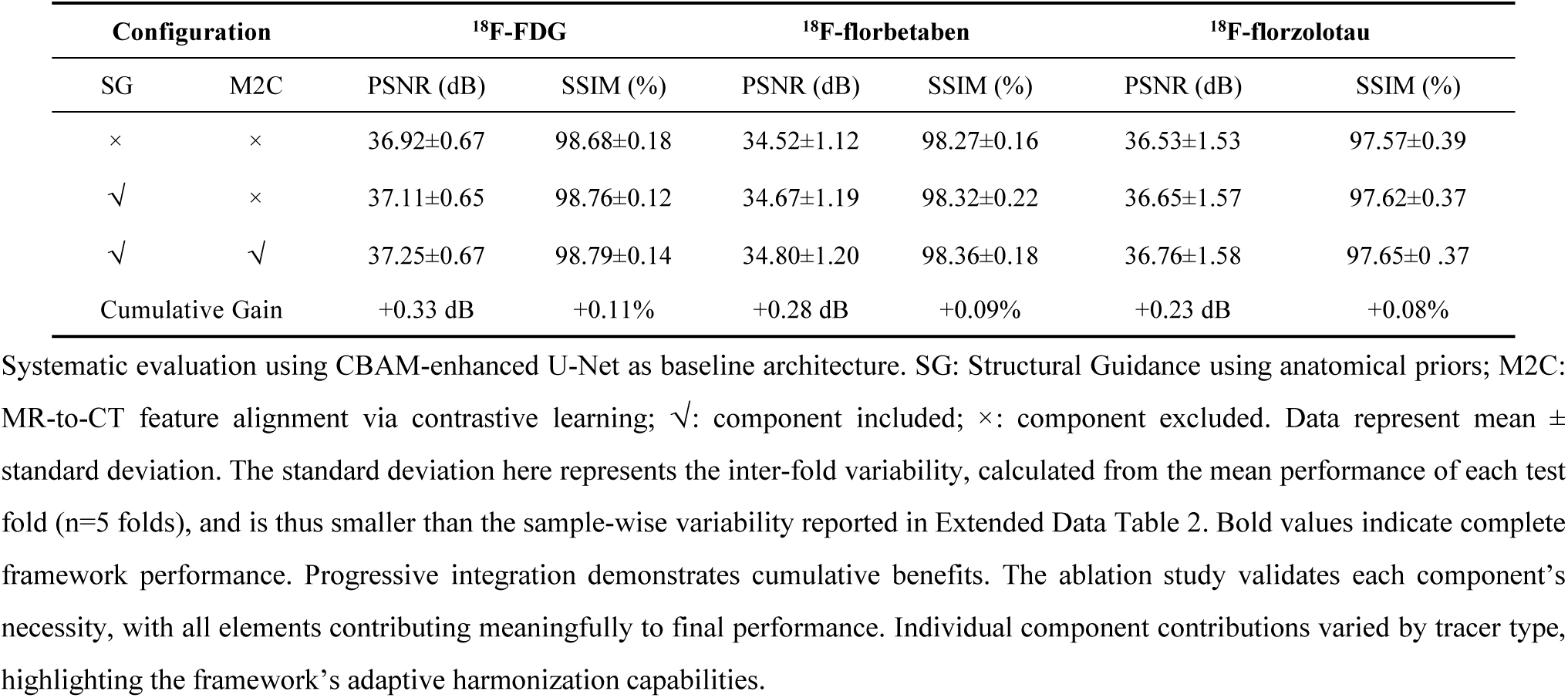
Systematic ablation study of framework architectural components

