## Supplementary materials for "A unified deep learning framework for cross-platform harmonization of multi-tracer PET quantification"

### Supplementary results section

#### Supplementary Table 1 | Regional bias analysis for normalized intensity measurements

| Tracer | Comparison | brain region | Bias and Agreement | | | |
|---|---|---|---|---|---|---|
| | | | Bias (%) | SD (%) | Upper LoA (%) | Lower LoA (%) |
| $^{18}$F-florbetaben | PET-MRI vs PET-CT | Frontal meta-ROI | -7.80 | 6.44 | 4.83 | -20.42 |
| | | Lateral parietal meta-ROI | -3.13 | 6.52 | 9.66 | -15.91 |
| | | Lateral temporal meta-ROI | -7.15 | 5.73 | 4.09 | -18.38 |
| | | Cerebellum | -16.18 | 6.03 | -4.36 | -28.00 |
| | | Centiloid meta-ROI | -7.06 | 5.69 | 4.09 | -18.20 |
| | Harmonized PET-MRI vs PET-CT | Frontal meta-ROI | 0.70 | 4.72 | 9.95 | -8.55 |
| | | Lateral parietal meta-ROI | 0.58 | 5.52 | 11.39 | -10.23 |
| | | Lateral temporal meta-ROI | 0.50 | 5.23 | 10.75 | -9.75 |
| | | Cerebellum | 0.10 | 5.96 | 11.79 | -11.59 |
| | | Centiloid meta-ROI | 0.39 | 4.33 | 8.87 | -8.09 |
| | PET-MRI vs Harmonized PET-MRI | Frontal meta-ROI | -8.50 | 3.48 | -1.68 | -15.33 |
| | | Lateral parietal meta-ROI | -3.71 | 3.29 | 2.73 | -10.16 |
| | | Lateral temporal meta-ROI | -7.65 | 2.59 | -2.58 | -12.72 |
| | | Cerebellum | -16.29 | 3.06 | -10.29 | -22.30 |
| | | Centiloid meta-ROI | -7.45 | 2.45 | -2.65 | -12.25 |
| $^{18}$F-florzolotau | PET-MRI vs PET-CT | Frontal meta-ROI | -13.61 | 10.02 | 6.02 | -33.24 |
| | | Lateral parietal meta-ROI | -8.25 | 9.75 | 10.86 | -27.37 |
| | | Lateral temporal meta-ROI | -15.01 | 10.29 | 5.15 | -35.17 |
| | | CerebellumGM | -21.61 | 10.03 | -1.95 | -41.27 |
| | | CenTauR meta-ROI | -11.55 | 10.39 | 8.82 | -31.92 |
| | Harmonized PET-MRI vs PET-CT | Frontal meta-ROI | -3.85 | 10.81 | 17.33 | -25.03 |
| | | Lateral parietal meta-ROI | -3.94 | 9.73 | 15.12 | -23.00 |
| | | Lateral temporal meta-ROI | -6.34 | 10.53 | 14.29 | -26.97 |
| | | CerebellumGM | -4.57 | 13.30 | 21.51 | -30.64 |
| | | CenTauR meta-ROI | -4.71 | 10.09 | 15.08 | -24.49 |
| | PET-MRI vs Harmonized PET-MRI | Frontal meta-ROI | -9.78 | 5.77 | 1.53 | -21.10 |
| | | Lateral parietal meta-ROI | -4.32 | 6.29 | 8.00 | -16.64 |
| | | Lateral temporal meta-ROI | -8.70 | 5.41 | 1.89 | -19.30 |
| | | CerebellumGM | -17.09 | 8.45 | -0.53 | -33.65 |
| | | CenTauR meta-ROI | -6.86 | 5.98 | 4.86 | -18.59 |
| $^{18}$F-FDG | PET-MRI vs PET-CT | Frontal meta-ROI | -3.47 | 5.29 | 6.90 | -13.84 |
| | | Lateral parietal meta-ROI | 2.73 | 4.57 | 11.68 | -6.23 |
| | | Lateral temporal meta-ROI | -3.28 | 4.53 | 5.59 | -12.16 |
| | | Global | -0.64 | 4.24 | 7.68 | -8.95 |
| | | Pons | -1.18 | 7.45 | 13.43 | -15.78 |
| | Harmonized | Frontal meta-ROI | -0.50 | 3.71 | 6.77 | -7.77 |

| | | | | | |
|---|---|---|---|---|---|
| PET-MRI vs | Lateral parietal meta-ROI | 0.92 | 3.96 | 8.69 | -6.85 |
| PET-CT | Lateral temporal meta-ROI | -0.72 | 3.78 | 6.68 | -8.13 |
| | Global | 0.05 | 3.60 | 7.10 | -7.00 |
| | Pons | -0.49 | 5.73 | 10.75 | -11.73 |
| PET-MRI vs | Frontal meta-ROI | -2.97 | 2.52 | 1.97 | -7.90 |
| Harmonized | Lateral parietal meta-ROI | 1.81 | 2.25 | 6.21 | -2.60 |
| PET-MRI | Lateral temporal meta-ROI | -2.56 | 2.44 | 2.22 | -7.35 |
| | Global | -0.68 | 2.07 | 3.37 | -4.74 |
| | Pons | -0.69 | 3.48 | 6.14 | -7.52 |

Comprehensive bias analysis showing percentage differences, standard deviations, and 95% limits of agreement (LoA) for normalized intensity measurements across all composite brain regions for three tracers. Three pairwise comparisons evaluated: (1) unharmonized PET-MRI vs PET-CT reference (baseline platform discrepancies), (2) harmonized PET-MRI vs PET-CT reference (residual differences post-harmonization), (3) unharmonized vs harmonized PET-MRI (direct harmonization effects). Harmonization substantially reduced regional biases: $^{18}$F-florbetaben to <1% (94% reduction), $^{18}$F-florzolotau to <7% (59-79% reduction), $^{18}$F-FDG to <1% (92% reduction). Positive bias values indicate higher uptake in first-listed modality; negative values indicate lower uptake. LoA, limits of agreement calculated as bias ± 1.96×SD. PET-CT serves as reference standard. CerebellumGM, cerebellar gray matter reference region for tau tracers; Centiloid meta-ROI, cortical composite for amyloid quantification; CenTauR meta-ROI, temporal composite for tau quantification. Data represent percentage differences calculated at subject level and averaged across participants. Region-specific visualization in main Fig. 3-5a.

### Supplementary Table 2 | Regional bias analysis for SUVR measurements

| Tracer | **Comparison** | **brain region** | **Bias and Agreement** | | | |
|---|---|---|---|---|---|---|
| | | | **Bias (%)** | **SD (%)** | **Upper LoA (%)** | **Lower LoA (%)** |
| $^{18}$F-florbetaben | PET-MRI vs | Frontal meta-ROI | 8.41 | 6.14 | 20.44 | -3.62 |
| | PET-CT | Lateral parietal meta-ROI | 13.07 | 6.14 | 25.12 | 1.03 |
| | | Lateral temporal meta-ROI | 9.06 | 4.77 | 18.42 | -0.29 |
| | | Centiloid meta-ROI | 7.78 | 5.35 | 18.26 | -2.69 |
| | Harmonized | Frontal meta-ROI | 0.60 | 4.10 | 8.64 | -7.44 |
| | PET-MRI vs | Lateral parietal meta-ROI | 0.48 | 4.26 | 8.83 | -7.86 |
| | PET-CT | Lateral temporal meta-ROI | 0.40 | 3.74 | 7.74 | -6.94 |
| | | Centiloid meta-ROI | -0.11 | 2.98 | 5.74 | -5.96 |
| | PET-MRI vs | Frontal meta-ROI | 7.81 | 4.03 | 15.72 | -0.09 |
| | Harmonized | Lateral parietal meta-ROI | 12.60 | 4.32 | 21.07 | 4.12 |
| | PET-MRI | Lateral temporal meta-ROI | 8.67 | 3.12 | 14.79 | 2.55 |
| | | Centiloid meta-ROI | 7.90 | 3.76 | 15.27 | 0.52 |
| $^{18}$F-florzolotau | PET-MRI vs | Frontal meta-ROI | 8.31 | 4.91 | 17.93 | -1.32 |
| | PET-CT | Lateral parietal meta-ROI | 13.62 | 5.05 | 23.51 | 3.72 |
| | | Lateral temporal meta-ROI | 9.28 | 4.68 | 18.44 | 0.11 |
| | | CenTauR meta-ROI | 10.10 | 6.43 | 22.71 | -2.51 |
| | Harmonized | Frontal meta-ROI | 0.81 | 5.86 | 12.29 | -10.68 |
| | PET-MRI vs | Lateral parietal meta-ROI | 0.51 | 6.18 | 12.62 | -11.61 |
| | PET-CT | Lateral temporal meta-ROI | -0.69 | 6.31 | 11.68 | -13.06 |
| | | CenTauR meta-ROI | -0.71 | 8.22 | 15.41 | -16.83 |
| | PET-MRI vs | Frontal meta-ROI | 7.50 | 6.52 | 20.27 | -5.28 |
| | Harmonized | Lateral parietal meta-ROI | 13.11 | 6.76 | 26.35 | -0.13 |
| | PET-MRI | Lateral temporal meta-ROI | 9.96 | 7.15 | 23.97 | -4.05 |
| | | CenTauR meta-ROI | 10.81 | 7.81 | 26.13 | -4.50 |
| $^{18}$F-FDG | PET-MRI vs | Frontal meta-ROI | -2.3 | 7.5 | 12.5 | -17.0 |
| | PET-CT | Lateral parietal meta-ROI | 3.9 | 7.3 | 18.2 | -10.4 |
| | | Lateral temporal meta-ROI | -2.1 | 6.3 | 10.2 | -14.4 |
| | | Global | 0.5 | 6.8 | 13.9 | -12.8 |
| | Harmonized | Frontal meta-ROI | -0.0 | 5.6 | 11.0 | -11.0 |
| | PET-MRI vs | Lateral parietal meta-ROI | 1.4 | 6.0 | 13.2 | -10.3 |
| | PET-CT | Lateral temporal meta-ROI | -0.2 | 4.9 | 9.5 | -9.9 |
| | | Global | 0.5 | 5.4 | 11.0 | -10.0 |
| | PET-MRI vs | Frontal meta-ROI | -2.3 | 3.4 | 4.4 | -9.0 |
| | Harmonized | Lateral parietal meta-ROI | 2.5 | 3.2 | 8.8 | -3.8 |
| | PET-MRI | Lateral temporal meta-ROI | -1.9 | 2.9 | 3.8 | -7.6 |
| | | Global | 0.0 | 3.1 | 6.1 | -6.1 |

Comprehensive bias analysis showing percentage differences, standard deviations, and 95% limits of agreement for SUVR measurements across composite brain regions for three tracers. SUVR calculated using tracer-specific reference regions: whole cerebellum for $^{18}$F-florbetaben, cerebellar gray matter for $^{18}$F-florzolotau, pons for $^{18}$F-FDG. Three pairwise comparisons as in

Extended Data Table 3. Harmonization reduced SUVR biases: $^{18}$F-florbetaben from 7.78-13.07% to -0.11-0.60%, $^{18}$F-florzolotau from 8.31-13.62% to -0.71-0.81%, $^{18}$F-FDG maintained low biases. Concordance between normalized intensity (Extended Data Table 4) and SUVR findings confirms robust harmonization independent of quantification metric. SUVR, standardized uptake value ratio; bias calculation and statistical methods identical to Extended Data Table 3. Reference regions: whole cerebellum ($^{18}$F-florbetaben), cerebellar gray matter ($^{18}$F-florzolotau), pons ($^{18}$F-FDG). CenTauR meta-ROI, temporal composite normalized to cerebellar gray matter; Centiloid meta-ROI, cortical composite normalized to whole cerebellum. Positive bias indicates higher SUVR in first-listed modality. Data represent percentage differences at subject level (participant counts as in Extended Data Table 3). Corresponding SUVR distributions and regression analyses in Extended Data Fig. 1.

### Supplementary Table 3 | Linear regression parameters for normalized intensity measurements

| Tracer | Brain Region | Equation | Slope | Intercept | *r* | $R^2$ | *P* Value | Significance |
|---|---|---|---|---|---|---|---|---|
| | Frontal meta-ROI | y = 0.920x + 0.042 | 0.92 | 0.04 | 0.97 | 0.95 | 2.26E-12 | *** |
| | Cingulate meta-ROI | y = 0.867x + 0.077 | 0.87 | 0.08 | 0.97 | 0.93 | 2.34E-11 | *** |
| | Lateral parietal meta-ROI | y = 0.905x + 0.051 | 0.90 | 0.05 | 0.96 | 0.92 | 8.00E-11 | *** |
| $^{18}$F-florbetaben | Lateral temporal meta-ROI | y = 0.877x + 0.061 | 0.88 | 0.06 | 0.96 | 0.93 | 4.83E-11 | *** |
| | Temporal meta-ROI | y = 0.812x + 0.086 | 0.81 | 0.09 | 0.91 | 0.82 | 9.27E-08 | *** |
| | Cerebellum | y = 0.727x + 0.118 | 0.73 | 0.12 | 0.87 | 0.75 | 1.57E-06 | *** |
| | Centiloid meta-ROI | y = 0.932x + 0.042 | 0.93 | 0.04 | 0.97 | 0.95 | 2.02E-12 | *** |
| | Frontal meta-ROI | y = 0.933x + 0.010 | 0.93 | 0.01 | 0.96 | 0.92 | 1.38E-10 | *** |
| | Cingulate meta-ROI | y = 0.950x + 0.003 | 0.95 | 0.00 | 0.96 | 0.93 | 4.00E-11 | *** |
| | Lateral parietal meta-ROI | y = 0.992x - 0.008 | 0.99 | -0.01 | 0.97 | 0.94 | 5.62E-12 | *** |
| $^{18}$F-florzolotau | Lateral temporal meta-ROI | y = 0.921x + 0.008 | 0.92 | 0.01 | 0.95 | 0.91 | 2.44E-10 | *** |
| | Temporal meta-ROI | y = 0.903x + 0.014 | 0.90 | 0.01 | 0.95 | 0.90 | 5.87E-10 | *** |
| | CerebellumGM | y = 0.851x + 0.024 | 0.85 | 0.02 | 0.97 | 0.94 | 1.55E-11 | *** |
| | CenTauR meta-ROI | y = 0.962x - 0.002 | 0.96 | 0.00 | 0.96 | 0.93 | 3.13E-11 | *** |
| | Frontal meta-ROI | y = 0.865x + 0.073 | 0.86 | 0.07 | 0.90 | 0.82 | 5.03E-08 | *** |
| | Parietal meta-ROI | y = 0.744x + 0.143 | 0.74 | 0.14 | 0.89 | 0.79 | 1.95E-07 | *** |
| | Temporal meta-ROI | y = 0.805x + 0.090 | 0.80 | 0.09 | 0.85 | 0.73 | 1.90E-06 | *** |
| $^{18}$F-FDG | Occipital meta-ROI | y = 0.726x + 0.157 | 0.73 | 0.16 | 0.91 | 0.84 | 1.65E-08 | *** |
| | Pons | y = 0.930x + 0.025 | 0.93 | 0.03 | 0.92 | 0.85 | 8.07E-09 | *** |
| | Global meta-ROI | y = 0.723x + 0.149 | 0.72 | 0.15 | 0.89 | 0.79 | 1.67E-07 | *** |

Linear regression parameters demonstrating quantitative relationships between harmonized PET-MRI and PET-CT reference across brain regions for three tracers. Each row represents regression of harmonized PET-MRI (dependent variable) against PET-CT reference (independent variable). Strong correlations combined with near-unity slopes confirm PET-CT-equivalent quantification while preserving inter-subject variability essential for clinical interpretation: $^{18}$F-florbetaben (*r*: 0.87-0.97, slopes: 0.73-0.93), $^{18}$F-florzolotau (*r*: 0.95-0.97, slopes: 0.85-0.99), $^{18}$F-FDG (*r*: 0.85-0.92, slopes: 0.72-0.93). All correlations achieved P<0.001, confirming statistically robust cross-platform concordance. *r*, Pearson correlation coefficient; $R^2$, coefficient of determination; *P* value from two-tailed significance test of correlation. Regression equation format: y = slope×x + intercept, where y represents harmonized PET-MRI and x represents PET-CT reference. Significance levels: ****P* < 0.001. Ideal harmonization defined as near-unity slopes (0.9-1.1) with high correlations (*r* > 0.90). CerebellumGM, cerebellar gray matter; CenTauR meta-ROI, temporal composite for tau quantification; Centiloid meta-ROI, cortical composite for amyloid quantification per Centiloid standard. Corresponding scatter plots in main Fig. 3-5b.

**Supplementary Table 4 | Linear regression parameters for SUVR measurements**

| Tracer | Brain Region | Equation | Slope | Intercept | r | R2 | P Value | Significance |
|---|---|---|---|---|---|---|---|---|
| $^{18}$F-florbetaben | Frontal meta-ROI | y = 0.923x + 0.094 | 0.92 | 0.09 | 0.97 | 0.95 | 1.72E-12 | *** |
| | Cingulate meta-ROI | y = 0.905x + 0.130 | 0.90 | 0.13 | 0.96 | 0.92 | 1.07E-10 | *** |
| | Lateral parietal meta-ROI | y = 0.925x + 0.094 | 0.93 | 0.09 | 0.97 | 0.94 | 5.19E-12 | *** |
| | Lateral temporal meta-ROI | y = 0.902x + 0.113 | 0.90 | 0.11 | 0.97 | 0.95 | 2.30E-12 | *** |
| | Temporal meta-ROI | y = 0.946x + 0.059 | 0.95 | 0.06 | 0.96 | 0.93 | 3.42E-11 | *** |
| | Centiloid meta-ROI | y = 0.930x + 0.087 | 0.93 | 0.09 | 0.99 | 0.97 | 1.17E-14 | *** |
| $^{18}$F-florzolotau | Frontal meta-ROI | y = 0.892x + 0.144 | 0.89 | 0.14 | 0.98 | 0.97 | 2.51E-13 | *** |
| | Cingulate meta-ROI | y = 0.851x + 0.204 | 0.85 | 0.20 | 0.98 | 0.96 | 2.21E-12 | *** |
| | Lateral parietal meta-ROI | y = 0.842x + 0.213 | 0.84 | 0.21 | 0.98 | 0.96 | 7.25E-13 | *** |
| | Lateral temporal meta-ROI | y = 0.860x + 0.179 | 0.86 | 0.18 | 0.98 | 0.96 | 2.82E-12 | *** |
| | Medial temporal meta-ROI | y = 0.862x + 0.180 | 0.86 | 0.18 | 0.97 | 0.93 | 8.72E-11 | *** |
| | CenTauR meta-ROI | y = 0.790x + 0.327 | 0.79 | 0.33 | 0.97 | 0.93 | 8.80E-11 | *** |
| $^{18}$F-FDG | Frontal meta-ROI | y = 0.885x + 0.169 | 0.88 | 0.17 | 0.92 | 0.85 | 6.09E-09 | *** |
| | Parietal meta-ROI | y = 0.938x + 0.110 | 0.94 | 0.11 | 0.91 | 0.83 | 2.28E-08 | *** |
| | Temporal meta-ROI | y = 0.869x + 0.163 | 0.87 | 0.16 | 0.91 | 0.83 | 2.38E-08 | *** |
| | Occipital meta-ROI | y = 0.953x + 0.083 | 0.95 | 0.08 | 0.92 | 0.85 | 8.92E-09 | *** |
| | Global meta-ROI | y = 0.910x + 0.135 | 0.91 | 0.13 | 0.92 | 0.84 | 1.32E-08 | *** |

Linear regression parameters for SUVR measurements demonstrating preserved quantitative relationships following harmonization. SUVR calculated using tracer-specific reference regions as detailed in Extended Data Table 4. Regression of harmonized PET-MRI SUVR (dependent variable) against PET-CT reference SUVR (independent variable). SUVR-based analyses achieved even stronger concordance than normalized intensity measurements: $^{18}$F-florbetaben (*r*: 0.96-0.99, slopes: 0.90-0.95), $^{18}$F-florzolotau (*r*: 0.97-0.98, slopes: 0.79-0.89), $^{18}$F-FDG (*r*: 0.91-0.92, slopes: 0.87-0.95). All correlations $P < 0.001$. Consistency between normalized intensity (Extended Data Table 6) and SUVR regression parameters confirms robust cross-platform calibration independent of quantification framework. SUVR, standardized uptake value ratio; statistical parameters and notation identical to Extended Data Table 6. Reference regions for normalization: whole cerebellum ($^{18}$F-florbetaben), cerebellar gray matter ($^{18}$F-florzolotau), pons ($^{18}$F-FDG). CenTauR, CenTauR temporal composite meta-ROI; Centiloid meta-ROI, Centiloid cortical composite meta-ROI. Regression equation: y = slope×x + intercept (y: harmonized PET-MRI SUVR, x: PET-CT reference SUVR). Significance: ***$P < 0.001$. Near-perfect concordance ($r > 0.96$ for amyloid/tau, $r > 0.91$ for metabolic imaging) confirms preserved inter-subject relationships essential for diagnostic classification. Corresponding SUVR distributions and scatter plots in Extended Data Fig. 1.

**Supplementary Table5 | Brain regions used for inter-regional correlation analysis**

| Region name | Brain lobe-abbreviation | Brain lobe | Brain lobe-abbreviation |
|---|---|---|---|
| ctx-bankssts | BSTS | Temporal | TC |
| ctx-caudalanteriorcingulate | CACG | Cingulate | CC |
| ctx-caudalmiddlefrontal | CMFG | Frontal | FC |
| ctx-cuneus | CU | Occipital | OC |
| ctx-entorhinal | EC | Temporal | TC |
| ctx-fusiform | FG | Temporal | TC |
| ctx-inferiorparietal | IPG | Parietal | PC |
| ctx-inferiortemporal | ITG | Temporal | TC |
| ctx-isthmuscingulate | ICG | Cingulate | CC |
| ctx-lateraloccipital | LOG | Occipital | OC |
| ctx-lateralorbitofrontal | LOFG | Frontal | FC |
| ctx-lingual | LG | Occipital | OC |
| ctx-medialorbitofrontal | MOFG | Frontal | FC |
| ctx-middletemporal | MTG | Temporal | TC |
| ctx-parahippocampal | PHIG | Temporal | TC |
| ctx-paracentral | PaCG | Frontal | FC |
| ctx-parsopercularis | POP | Frontal | FC |
| ctx-parsorbitalis | POR | Frontal | FC |
| ctx-parstriangularis | PTR | Frontal | FC |
| ctx-pericalcarine | PCAL | Occipital | OC |
| ctx-postcentral | PstCG | Parietal | PC |
| ctx-posteriorcingulate | PCG | Cingulate | CC |
| ctx-precentral | PreCG | Frontal | FC |
| ctx-precuneus | PCU | Parietal | PC |
| ctx-rostralanteriorcingulate | RACG | Cingulate | CC |
| ctx-rostralmiddlefrontal | RMFG | Frontal | FC |
| ctx-superiorfrontal | SFG | Frontal | FC |
| ctx-superiorparietal | SPG | Parietal | PC |
| ctx-superiortemporal | STG | Temporal | TC |
| ctx-supramarginal | SMG | Parietal | PC |
| ctx-frontalpole | FP | Frontal | FC |
| ctx-temporalpole | TP | Temporal | TC |
| ctx-transversetemporal | TTG | Temporal | TC |

**Supplementary methods: Multi-vendor scanner specifications and acquisition parameters**

**Development Cohort (HS-HQ)**

*PET-MRI Acquisition (United Imaging uPMR790):*

PET-MRI scans were acquired on a 3T PET/MR system (uPMR790, United Imaging Healthcare, Shanghai, China). Static PET data were reconstructed using all list mode events with ordered subset expectation maximization algorithm (4 iterations, 20 subsets). Acquisition parameters: field of view (FOV) = 300 × 300 $mm^2$, matrix size = 150 × 150, voxel size = 2.0 × 2.0 × 2.0 $mm^3$. Data were reconstructed after correction for randoms, dead time, scatter, and attenuation.

For MRI, a 3D Dixon sequence was acquired for attenuation correction, and a T1-weighted MR scan was simultaneously acquired using the following parameters: repetition time = 7200 ms, echo time = 3.0 ms, flip angle = 10°, acquisition matrix = 256 × 329, in-plane resolution = 1 mm × 1 mm, slice thickness = 1 mm, and 176 slices.

*PET-CT Acquisition (United Imaging uMI780):*

PET-CT was conducted using a uMI780 scanner (United Imaging Healthcare, Shanghai, China). A 20-minute PET acquisition with low-dose CT scan for attenuation correction was performed. Static PET data were reconstructed using all list mode events with ordered subset expectation maximization algorithm (4 iterations, 20 subsets). Acquisition parameters: FOV = 300 × 300 $mm^2$, matrix size = 150 × 150, voxel size = 2.0 × 2.0 × 2.0 $mm^3$.

**External Validation Cohort (AH-PH)**

*PET-MRI Acquisition (Siemens Biograph mMR):*

PET-MRI imaging was performed using a Siemens Biograph mMR scanner. Static PET data were reconstructed using all list mode events with ordered subset expectation maximization algorithm (8 iterations, 21 subsets). Acquisition parameters: FOV = 300 × 300 $mm^2$, matrix size = 150 × 150, voxel size = 2.0 × 2.0 × 2.0 $mm^3$. Data were reconstructed after correction for randoms, dead time, scatter, and attenuation.

For MRI, a 3D Dixon sequence was acquired for attenuation correction, and a T1-weighted MR scan was simultaneously acquired using the following parameters: repetition time = 2000 ms, echo time = 3.24 ms, flip angle = 7°, acquisition matrix = 256 × 256, slice thickness = 1 mm, and 192 slices.

*PET-CT Acquisition (Siemens Biograph Vision 600):*

PET-CT imaging was performed using a Siemens Biograph Vision 600 scanner. Following radiotracer injection, a 20-minute static PET acquisition was conducted in supine position with head-first orientation. Low-dose CT scans (120 kVp) were acquired for attenuation correction using the manufacturer's standard brain protocol. PET data were reconstructed using ordered subset expectation maximization (OSEM) algorithm with point spread function and time-of-flight corrections (PSF+TOF, 10 iterations, 5 subsets).

**Clinical Validation Cohort**

*XWH Site - GE SIGNA PET/MR 3.0T:*

PET-MRI scans were acquired on a GE SIGNA PET/MR 3.0T system. Static PET data were reconstructed using vendor-specific algorithm. Data were reconstructed after correction for randoms, dead time, scatter, and attenuation. For MRI, a 3D Dixon sequence was acquired for attenuation correction, and a T1-weighted MR scan was simultaneously acquired using the following parameters: repetition time = 8488 ms, echo time = 3.25 ms, flip angle = 15°, acquisition matrix = 256 × 256, slice thickness = 1 mm, and 188 slices.

*XWH Site - United Imaging uPMR790:*

PET-MRI scans were acquired on a United Imaging uPMR790 3T PET/MR system. Static PET data were reconstructed using all list mode events with 3D iterative TOF PSF (Time-of-Flight with Point Spread Function correction). Acquisition parameters: FOV = 250 × 250 $mm^2$, matrix size = 150 × 150, voxel size = 1.5 × 1.5 × 1.5 $mm^3$. Data were reconstructed after correction for randoms, dead time, scatter, and attenuation. For MRI, a 3D Dixon sequence was acquired for attenuation correction, and a T1-weighted MR scan was simultaneously acquired using the following parameters: repetition time = 7200 ms, echo time = 3.0 ms, flip angle = 10°, acquisition matrix = 256 × 256, slice thickness = 1 mm, and 192 slices.

*HS-Gamma Site - Siemens Biograph mCT Flow:*

PET-CT was conducted using a Biograph mCT Flow scanner (Siemens, Erlangen, Germany). A 20-minute PET acquisition with low-dose CT scan for attenuation correction was performed. PET images were reconstructed using filtered back-projection algorithm with the following parameters: image size of 256 × 256, zoom factor of 2.00, Gaussian filter with full width at half maximum (FWHM) of 3.5 mm, and corrections for decay, normalization, dead time, attenuation, scatter, and random coincidences.
